# Estimation of biological age acceleration based on NMR metabolomics and other risk factors in the Estonian Biobank

**DOI:** 10.1101/2025.08.17.25333849

**Authors:** Māra Delesa-Vēliņa, Jaanika Kronberg, Estonian Biobank Research Team, Krista Fischer

## Abstract

A high-throughput platform of nuclear magnetic resonance (NMR) spectroscopy blood metabolite data from 30-90 years old participants of Estonian Biobank (*N* = 150, 023) is used to develop and validate a biomarker score for mortality and the corresponding biological age estimate. We define biological age as the age where an individual’s survival probability given their covariate profile matches the survival probability of an average individual in the population. We estimate the survival probabilities parametrically based on Gompertz model using the newly developed metabolite biomarker score and common risk factors. The resulting biological age estimate, *SurvMetaboAge*, is a powerful predictor of both overall and cause-specific mortality. One year of biological age acceleration (BAA, difference between individual’s biological and chronological age) is associated with 17% (95% CI 15%–18%) and 12% (95% CI 11%–13%) increase of hazards for overall mortality in the validation set for men and women, respectively. An appealing interpretation of BAA can be provided, demonstrating that it is an easily interpretable predictor of mortality encompassing metabolite profile and common risk factors in a single measure with a potential in risk stratification and risk communication.

## 1 Introduction

It is well known that the chronological age fails to capture the heterogeneity in health status among older adults [28], thus there is an interest to study alternative, i.e., biological, age measures. Biomarker of ageing, as first defined by *Baker et al.* [1], is a biological parameter that predicts functional capability at a later age better than chronological age alone. Accurate biomarkers of ageing and biological age measures can help addressing the economic and healthcare challenges posed by increased longevity and the prevalence of age-related diseases [22].

A variety of markers of biological ageing have been proposed based on the biological systems that alter with age, such as epigenetic (DNA methylation, DNAm) changes, proteomic changes, inflammatory and immune pathways, neuroimaging, microbiome-associated changes and other, see [20], [43] and [32] for an overview. Metabolomics, a profiling of small molecules involved in metabolic pathways and carried out via, for example, nuclear magnetic resonance (NMR) spectroscopy, have shown promise in developing new biomarkers of ageing [41]. A high throughput NMR metabolomics platform (Nightingale Health Ltd., Helsinki, Finland) [52] has been used to develop biomarker composite scores that are strongly associated with dimensions of ageing, including mortality and morbidity [9], [7], [53], and biological age estimates [47], [35], [26]. In a recent study NMR blood metabolomics data of a large cohort of three national biobanks was used to develop and validate scores for the prediction of 12 common morbidity-causing diseases [38]. For most of the diseases these scores were more strongly associated with disease onset than the polygenic risk scores.

In our previous research in the Estonian Biobank (EstBB), four NMR metabolite biomarkers (analytical sample size *n* = 9, 842) were found to be independently associated with mortality [9], while in a later meta-analysis where EstBB also took part [7], 14 significant metabolites were identified. Given the total number of participants with NMR metabolomics data (EstBB) now exceeds 200, 000, the aim of our current study is to develop an updated NMR-metabolites based score for all-cause mortality. In addition, we aim to develop a corresponding biological age estimate encompassing metabolite profile and common risk factors.

From a methodological point of view, biological age estimation methods (also called biological age clocks) can be divided into several generations [6]. The first generation clocks are based on the association between candidate markers and chronological age using (penalized) regression models [11], [14], [47], [42] or more recently various machine learning methods [35]. It has been hypothesized that the predicted age can serve as a measure of biological age, while the difference between the predicted age and chronological age (biological age acceleration) can provide more insight into the process of ageing independent of the chronological age [14], [11].

The approach of using age as a dependent variable is, however, problematic. One can note that the regression approach assumes that the biological age is defined as *E*(*A_i_|*X_i_), where *A_i_* is individual’s chronological age and X_i_ a vector of biomarkers. Thus the best biological age estimators are the ones that predict individual’s age as precisely as possible. This, however does not necessarily imply that individuals whose estimated biological age is higher than their chronological age, have a higher risk of diseases and a shorter lifespan. To understand, suppose that a subset of individuals in the analysed cohort are exposed to a risk factor, such as smoking, for instance. Additionally, suppose that smokers tend to be younger, in general. If some biomarkers in X_i_ are sensitive to smoking, this may lead to a finding that smokers are “biologically younger”. Since in the underlying true data-generating model the biomarkers are actually dependent variables, depending on both age and various other covariates, adjustment for smoking and other known risk factor in the model for age would not remove such contradictory findings. Moreover, regression models often produce biased estimates where the age is overestimated for the younger individuals and underestimated for the older individuals, and thus require a correction [25].

An alternative is to define the biological age in a manner that it is directly related to the underlying risk level, based on survival analysis model. The second generation biological age clocks have thus been developed by associating the biomarkers of ageing to time to mortality or time to disease onset. First such clocks have been developed in the area of epigenetic clocks [27], [29]. Recently, second generation clocks based on blood biomarkers and various phenotypic variables [5], [40] as well as clocks based on NMR metabolomics [44], [26] have appeared.

We follow the approach of the second generation biological age clocks and develop a biological age estimate in two stages. In the first stage, we create an NMR metabolite biomarker score by stepwise Cox proportional hazards regression for all-cause mortality. In the second stage, we define the metabolome-survival-based biological age *MetaboSurvAge* as the age where individual’s current survival probability given their covariate profile equals the survival probability of an average individual in the study population. Similarly as in [27], we model the survival distribution parametrically through Gompertz distribution and use the newly developed NMR metabolite score and common risk factors as covariates.

Our results indicate that the NMR metabolite biomarker score stratifies the population survival risk well in both development and validation cohorts. Additionally, the resulting biological age acceleration, defined as the difference between individual’s *MetaboSurvAge* and their chronological age, is a strong predictor for the short-term mortality risk and outperforms common risk factors or NMR metabolite biomarker score alone. We also provide an explication on how individual’s covariate values contribute to his or her biological age acceleration in time units of years.

## 2 Results

### 2.1 Sample characteristics

Estonian Biobank is a large population biobank where participants have been recruited in two waves (cohorts): the first, 2002–2017 (mostly up to 2010) comprises about 50,000 participants, while the second, 2018 onwards (mostly up to 2019) comprises about 150,000 participants. There are notable differences between the two cohorts, such as follow-up length, collection protocols, distribution of baseline demographic factors and chronic disease prevalence; for a detailed overview on EstBB, see [34]. The cohort differences pose a challenge to survival modelling: the two cohorts cannot be analysed together directly. However, when analysed separately, they produce models that lead to different predicted risk levels for the same individual if used for out-of-sample predictions. Instead of either making a choice between the two cohorts or producing average risk estimates based on the two, we use the data differences for model validation: we develop the survival model on the first cohort with the longest follow-up time, and employ the second cohort for model validation.

The sample characteristics are presented in Table 1. After selecting 30–90 years old participants and excluding cases with missing observations (see Methods and Supplementary Material A for details), the analytical sample size comprised 150,007 participants, 31,358 and 118,649 in discovery and validation cohorts, respectively. The mean age at recruitment was 51.3 (SD 13.9) and 49.9 (SD 13.2) in the discovery and validation cohorts. The mortality data was obtained from the Estonian Death Registry as on April 2024. Among the participants surviving to the end of the study, the mean follow-up time was 14.4 years (range 0.9 to 20.4) and 5.3 years (range 1.6 to 6.3) in the discovery and validation cohorts.

**Table 1:** Characteristics of the two EstBB recruitment cohorts used to define the discovery (2002–2017) and validation (2018 onwards) sets.

|  | **Discovery (2002-2017)**<br>N=31,358 |  | **Validation (2018 onwards)**<br>N=118,649 |  |
| --- | --- | --- | --- | --- |
|  | Male<br>9,763 (31.1%)<sup>1</sup> | Female<br>21,595 (68.9%)<sup>1</sup> | Male<br>41,219 (34.7%)<sup>1</sup> | Female<br>77,430 (65.3%)<sup>1</sup> |
| **Baseline** |  |  |  |  |
| Age | 51.5 (14.1) | 51.2 (13.8) | 48.7 (13.0) | 50.6 (13.3) |
| Age distribution |  |  |  |  |
| 30–44 | 3,563 (36.5%) | 7,998 (37.0%) | 18,192 (44.1%) | 29,362 (37.9%) |
| 45–64 | 4,139 (42.4%) | 9,494 (44.0%) | 17,340 (42.1%) | 34,949 (45.1%) |
| 65–90 | 2,061 (21.1%) | 4,103 (19.0%) | 5,687 (13.8%) | 13,119 (16.9%) |
| Education level |  |  |  |  |
| Primary | 1,948 (20.0%) | 2,985 (13.8%) | 4,480 (11.0%) | 5,182 (6.8%) |
| Secondary | 5,194 (53.2%) | 11,740 (54.4%) | 19,615 (48.0%) | 33,327 (43.5%) |
| Tertiary | 2,619 (26.8%) | 6,867 (31.8%) | 16,757 (41.0%) | 38,079 (49.7%) |
| Missing | 2 | 3 | 367 | 842 |
| Current smoker | 3,432 (35.2%) | 4,220 (19.5%) | 8,382 (20.3%) | 10,740 (13.9%) |
| BMI | 27.4 (4.6) | 27.2 (5.6) | 27.6 (4.4) | 26.3 (5.4) |
| Missing | 62 | 109 | 3,937 | 5,985 |
| BMI > 35 | 570 (5.8%) | 2,049 (9.5%) | 2,271 (5.5%) | 5,133 (6.6%) |
| Prevalent chronic disease |  |  |  |  |
| Diabetes | 857 (8.8%) | 1,547 (7.2%) | 2,969 (7.2%) | 4,624 (6.0%) |
| Cardiovascular | 2,268 (23.2%) | 3,760 (17.4%) | 3,806 (9.2%) | 5,793 (7.5%) |
| Cancer | 724 (7.4%) | 1,423 (6.6%) | 2,791 (6.8%) | 5,596 (7.2%) |
| Statin use | 576 (5.9%) | 927 (4.3%) | 3,468 (8.4%) | 5,229 (6.8%) |
| **Outcome** |  |  |  |  |
| Years of follow-up | 12.5 (4.8) | 13.7 (4.2) | 5.18 (0.74) | 5.24 (0.65) |
| Total deaths | 2,537 (26.0%) | 3,015 (14.0%) | 1,182 (2.9%) | 1,156 (1.5%) |
| Death within 10 years | 1,629 (16.7%) | 1,740 (8.1%) | 1,182 (2.9%) | 1,156 (1.5%) |
| Death cause |  |  |  |  |
| Cancer | 701 (27.6%) | 814 (27.0%) | 311 (35.6%) | 356 (43.1%) |
| Cardiovascular | 1,177 (46.4%) | 1,543 (51.2%) | 260 (29.7%) | 224 (27.1%) |
| External | 170 (6.7%) | 108 (3.6%) | 96 (11.0%) | 50 (6.1%) |
| Other | 489 (19.3%) | 550 (18.2%) | 207 (23.7%) | 196 (23.7%) |

<sup>1</sup>n (%); mean (SD)

### 2.2 Effects of biomarkers on mortality

From 235 NMR metabolite biomarkers used in the analysis, 193 were univariately associated with the 10-year mortality after multiple testing correction (Supplementary Figures 4–5 and Supplementary Data 1). Among the statistically significant associations, the ten largest protective effects for mortality were observed for various small and medium sized high density lipoprotein measures, the total amount of phospholipids, and amino acids albumin and leucine. The ten largest risk effects corresponded to various larger size lipoprotein measures, HDL particle size, the ratio of omega-6 to omega-3 fatty acids, and an inflammation marker glycoprotein acetyls (GlycA). It is well known that NMR metabolite measures are highly correlated, thus Cox proportional hazards model with stepwise selection was used to select 17 independently significant metabolite biomarkers, the respective hazard ratio estimates being presented in Figure 1a. The three largest risk effects adjusted for other metabolites correspond to the ratio of docosahexaenoic acid to total fatty acids (DHA%), ratio of omega-6 fatty acids to omega-3 fatty acids, and GlycA concentration, while the three largest protective effects correspond to the degree of unsaturation of fatty acids, branched-chain amino acid valine concentration, and esterified cholesterol concentration in small size (average diameter 8.7 nm) high-density lipoprotein particles (S HDL CE).

**Figure 1:**
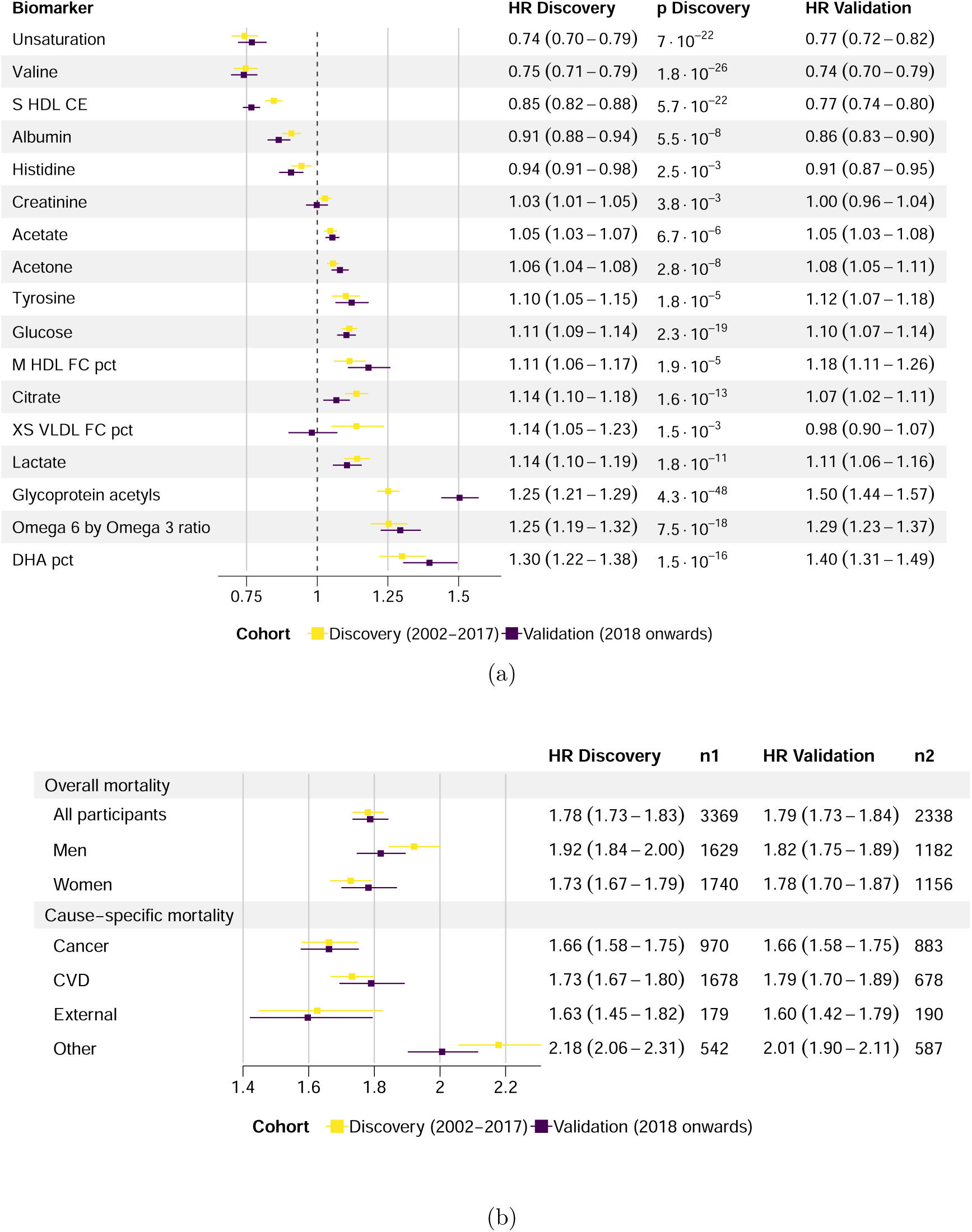
Hazard ratio (HR) estimates (with 95% confidence intervals) associated with 10-year mortality, discovery and validation cohorts. (a) Per SD change in metabolite biomarker value conditional on other metabolite values in the multivariate Cox model with 17 stepwise-selected biomarkers. (b) Per SD change in total biomarker score, for various subgroups and death causes. *n*1, *n*2 - number of outcomes in discovery and validation sets. XS, S, M – lipoprotein size subclass based on the lipoprotein particle diameter; HDL – high density lipoprotein; VLDL – very low density lipoprotein; CE – esterified cholesterol; FC – free cholesterol; pct – percentage measure; CVD – cardiovascular disease.

Metabolite effect directions were consistent in the unadjusted (univariate) case and when adjusted for all the other metabolites (multivariate) case, with two exceptions, DHA percentage among fatty acids (DHA%) and percentage of free cholesterol in medium size (average diameter 10.9 nm) high-density lipoprotein particles (*M HDL FC pct*). To explore these relationships, as a sensitivity analysis we estimated Cox models with all the combinations of variables among the 17 selected metabolites that were the most closely related to the metabolites demonstrating the adverse effects, see Supplementary Note B. First, we explored DHA% together with the other two metabolites describing fatty acid content (Unsaturation index and Omega 6 to 3 ratio). Second, we explored *M HDL FC pct* together with the other two variables describing lipid composition, *XS VLDL FC pct* and *S HDL CE*.

To evaluate the consistency of biomarker effects, the selected 17 biomarkers were used to retrain the Cox model in the validation set and for cause-specific mortality in discovery set. The direction and importance of the effects in the validation set were mostly consistent with those in the discovery set (Figure 1a), except for creatinine and *XS VLDL FC pct* not being significant in the validation cohort. Interestingly, glycoprotein acetyls had a markedly stronger association with the mortality in the validation set when compared to the discovery set. Regarding the cause-specific mortality, the effect directions are in general consistent across the mortality causes, with few exceptions (Supplementary Figure 7). First, death from the external factors (accidents etc.), shows significant associations with only some of the metabolites (Unsaturation index, Valine, Acetate, *M HDL FC pct*, GlycA, DHA%), while demonstrating opposite effects than other death causes for the citrate and being insignificant for others. Histidine did not appear to be significantly associated with any of the death categories, while tyrosine was only significant for the other death causes category, and creatinine only significant for the cancer category. Lack of significance could be partly explained by a smaller power due to fewer cases in the stratified analysis.

Hazard ratio estimates per one SD change in NMR metabolite biomarker score associated with overall and cause-specific 10-year mortality are presented in Figure 1b. The score was strongly associated with mortality both in discovery (HR 1.78 per SD, 95% CI 1.73–1.83) and validation (HR 1.79 per SD, 95% CI 1.73–1.84) sets. The association was stronger for men, especially in the discovery cohort. The score was also associated with the cause-specific mortality, the largest effect (HR 2.01 per SD, 95% CI 1.90–2.11 in the validation set) being observed for the other death cause category (many cases being related to chronic metabolic, liver, kidney and pulmonary conditions), followed by cardiovascular, cancer and external death cause categories.

NMR score stratified the mortality risk well for both males and females in the validation cohort, especially for the 1% highest risk strata (Figure 2). For example, the cumulative probability of death at age 60 at the highest 1% risk NMR score strata, 99%–100%, when compared with the 95–99% strata was 2.8 times higher for men (70.2%, 95% CI 54.4%–80.6% versus 24.7%, 95% CI 17.5%–31.3%) and 3.8 times higher for women (38.0%, 95% CI 22.7–50.3% versus 9.9%, 95% CI 6.3%–13.4%). The risk stratification capability of NMR was retained in different age groups (Supplementary Figures 9 and 10).

**Figure 2:**
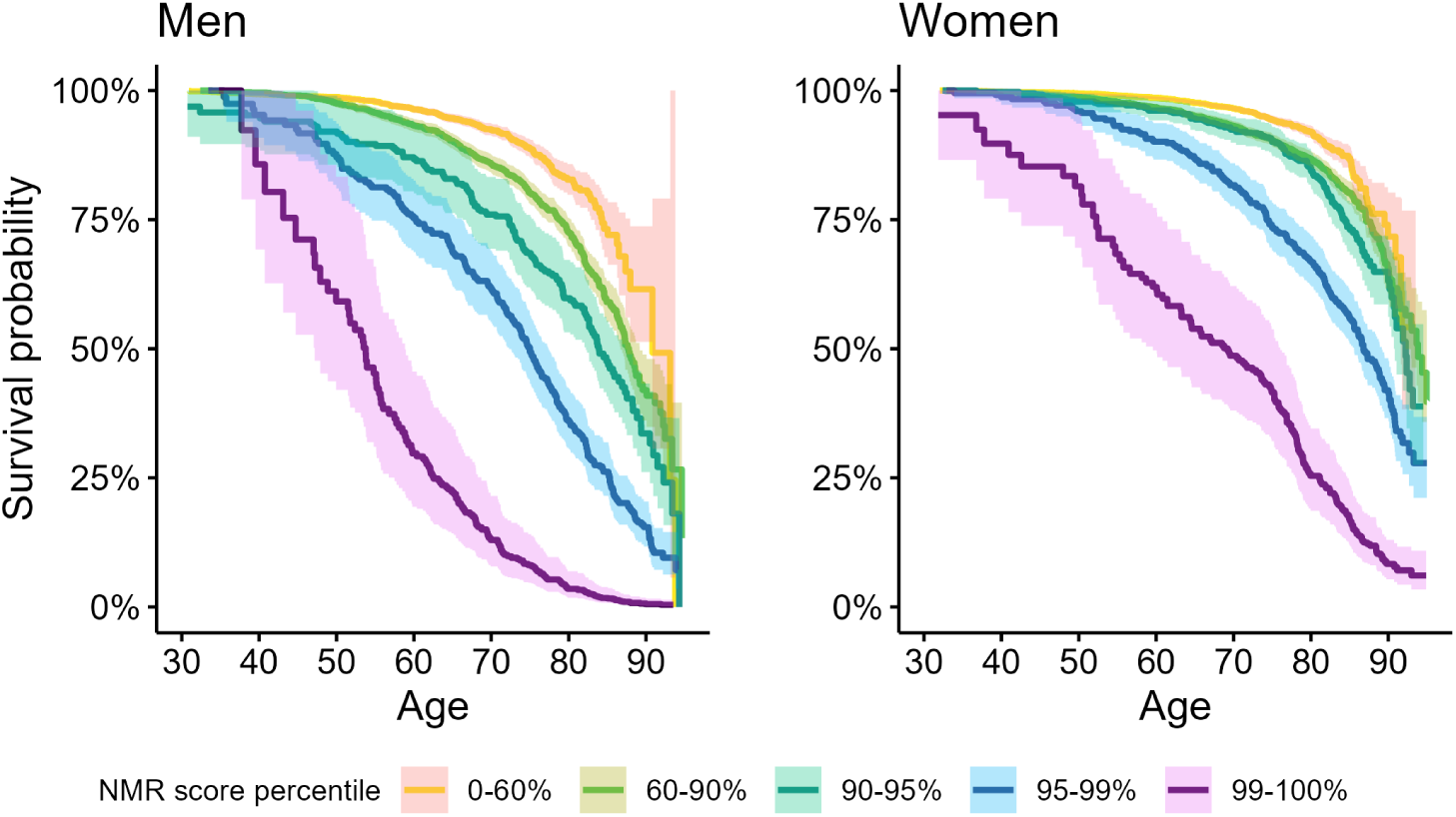
Estimates of overall mortality risk in validation set, stratified by the percentiles of the NMR score. The shaded regions represent 95% point-wise confidence intervals of the survival estimates. Left: men, right: women.

### 2.3 Survival- and metabolite-based biological age

We acquire the survival- and metabolite-based biological age estimate, *SurvMetaboAge*, by equating the unconditional survival function with the the survival function conditional on the standardized NMR score, common risk factors (BMI, education level, smoking) and prevalent chronic disease (cancer, diabetes and cardiovascular disease). *SurvMetaboAge* model parameter estimates are presented in Supplementary Table 4, the scatter plots of the biological age estimates with respect to the chronological age are presented in Figure 3. Biological age is highly correlated with the chronological age (Pearson’s *r* = 0.932, *p <* 0.001 and *r* = 0.938, *p <* 0.001 in discovery and validation cohorts, respectively). Biological age is approximately symmetrically distributed around the chronological age, although there are more extremes of biological age being higher than the chronological age (positive biological age acceleration), due to the fact that the NMR biomarker score is positively skewed in the dataset (Supplementary Figure 8). The average biological age (red dashed line) in each of the chronological age groups is slightly lower than the chronological age except for ages above 80 (85 in the validation set). In discovery set, the median biological age acceleration (BAA) is *−*2.9, IQR (*−*5.9, 0.7), 95% of estimates (*−*11.4, 9.6) years. In validation set, the median biological age acceleration is *−*3.5, IQR (*−*6.8*, −*0.6), 95% of estimates (*−*12.2, 7.0) years. We also compared the biological age distribution between the participants being alive and participants deceased at the end of the study grouped by their chronological age (Figure 4, validation cohort). The biological age estimates of the alive participants are below those of the deceased participants for all ages above 38, demonstrating good consistency with the survival status.

**Figure 3:**
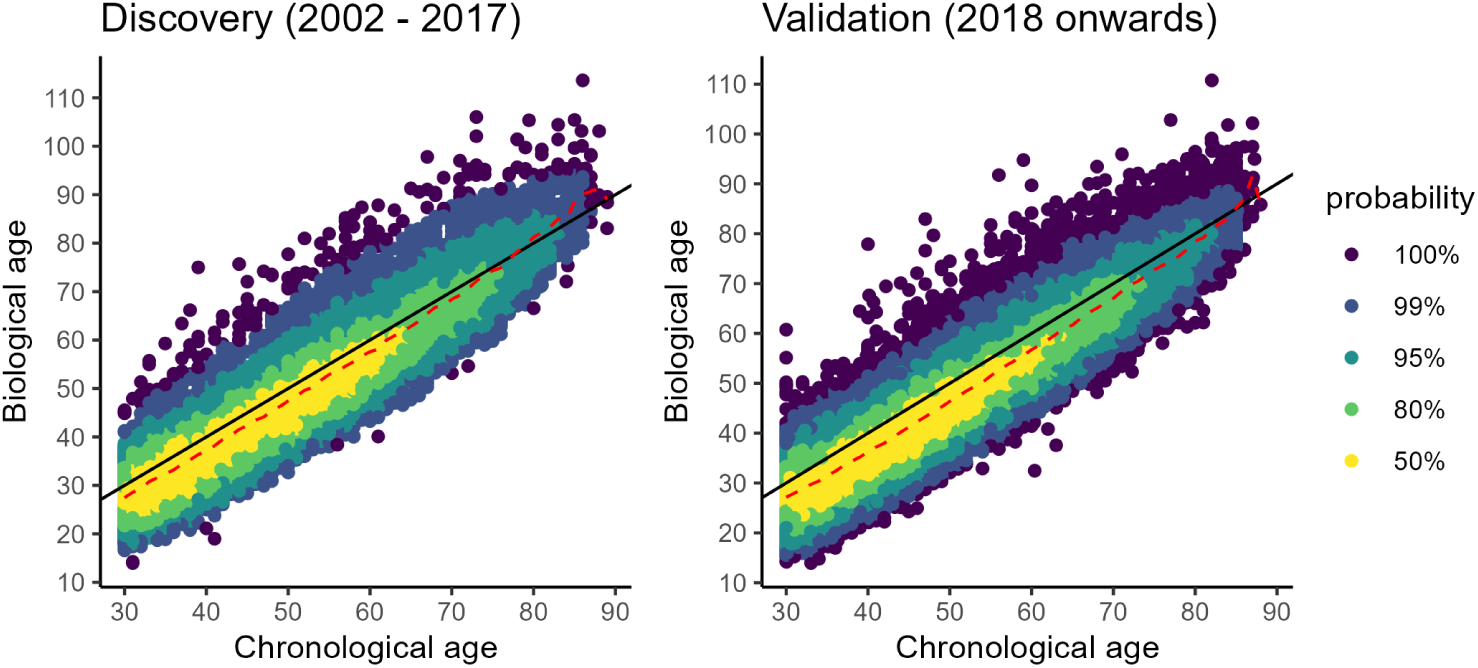
Association between the biological age estimate and the chronological age in discovery and validation sets. The black line is the diagonal where the chronological and biological ages are equal. The red dashed line represents the mean biological age in each of the chronological age groups split by age in full years. Colours represent the probabilities of 2-dimensional density estimates based on highest density regions.

**Figure 4:**
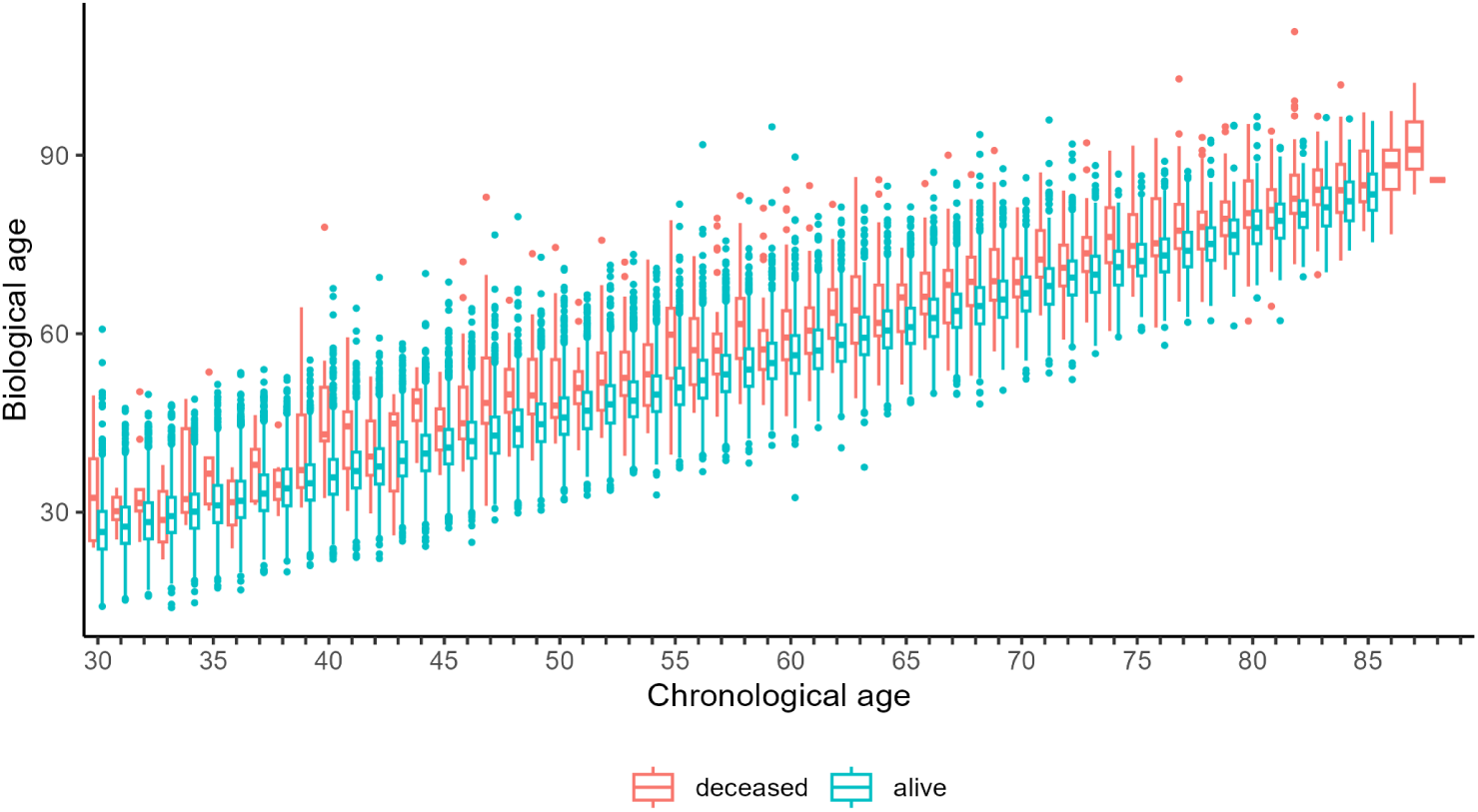
Biological age distribution compared between the participants being alive and deceased at the end of the study grouped by the chronological age, validation cohort.

We evaluate the predictive ability of the biological age acceleration by employing it in Cox proportional hazards model for overall mortality with age as a timescale (Table 2). One year of biological age acceleration is associated with HR 1.17 and 1.12 for men and women, respectively, in the validation set. One SD increase in the biological age acceleration is associated with HR of 1.93 and 1.94 for men and women, respectively. Note that these hazard ratios are larger than the hazard ratios associated with one SD increase of NMR metabolite score (1.82 and 1.78 for men and women, respectively, Supplementary Table 3).

**Table 2:** Hazard ratios (HR) associated with the biological age acceleration (BAA, difference between the individual’s biological age estimate, *SurvMetaboAge*, and their chronological age) in the Cox proportional hazards model for overall mortality, age as a timescale, validation cohort.

Next, we calculate Harell’s C-statistic to evaluate the capacity of the risk score based on biological age estimate in discriminating among subjects with different event times, and compare it to other potential risk scores (Figure 5). As we are using age as a timescale, chronological age is not informative in our model and corresponds to Cox model with an intercept only (C-statistic 0.5). NMR metabolite score alone exceeds the discrimination ability of common risk factors. NMR metabolite score performance is similar to that of common risk factors and prevalent chronic disease information combined, which might raise a speculation that NMR metabolite score reflects similar information as the rest of the predictors. Nevertheless, combining all the aforementioned covariates with NMR score further increases the C-statistic to 0.719 and 0.702 for men and women, respectively. Finally, practically the same predictive ability can be attained by using biological age acceleration as a single predictor, C-statistic 0.720 and 0.693 for men and women, respectively. It confirms that the biological age acceleration derived from *SurvMetaboAge* is a powerful predictor of mortality encompassing all the information of the predictors considered in a from of a single, easily interpretable measure. Note that there are some differences between the sexes: NMR metabolite score and common risk factors are more predictive for men, while information on prevalent disease is more predictive for women. Among the prevalent conditions, cancer is the most informative for the survival, followed by diabetes and finally cardiovascular conditions.

**Figure 5:**
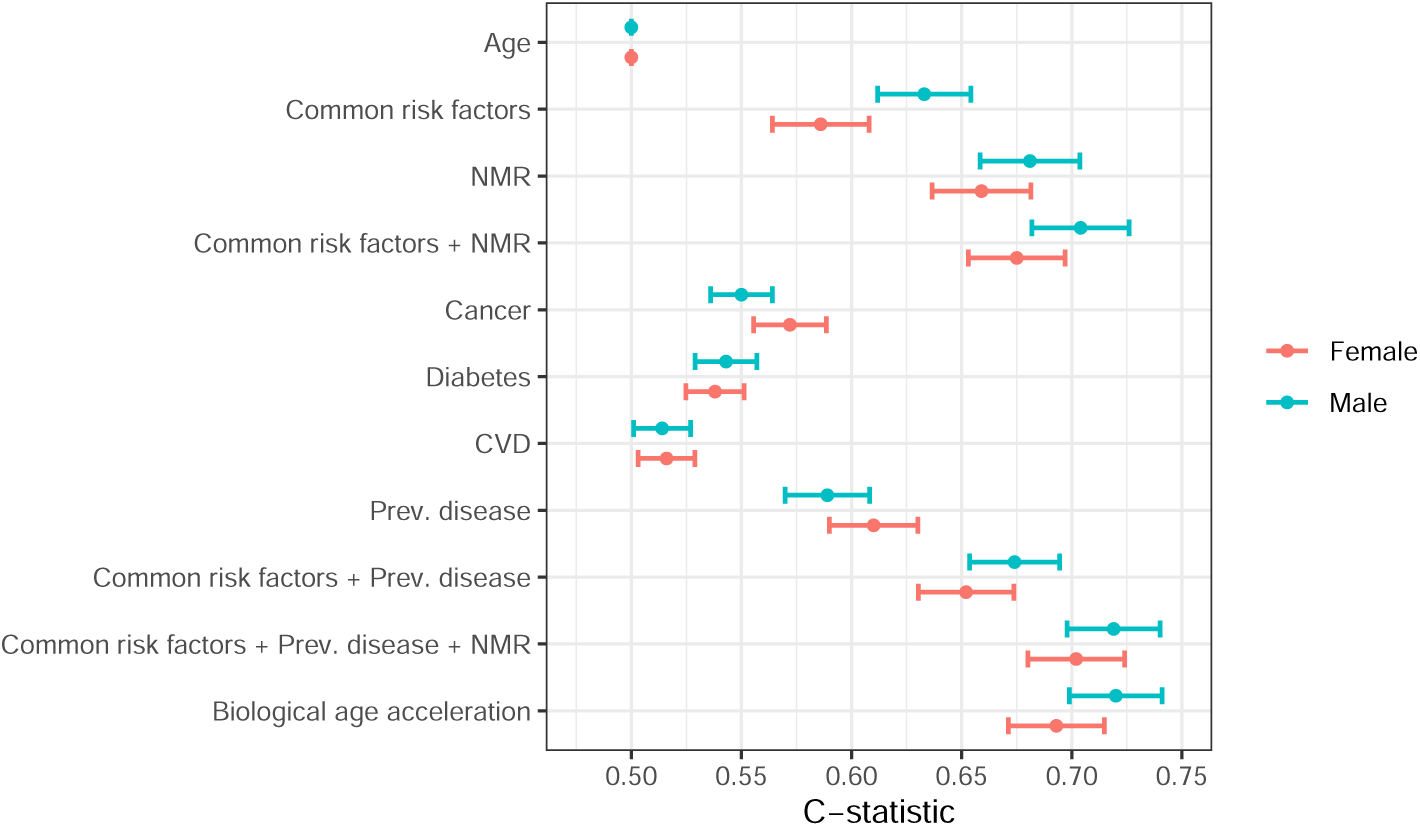
C-statistics of Cox proportional hazards model for overall survival in the validation cohort, age as a timescale, various covariate combinations. Models are evaluated separately for males and females. NMR: standardized NMR metabolite biomarker score; Common risk factors: BMI, education level, current smoking; CVD: cardiovascular disease; Prev. disease: combination of three covariates for prevalent disease status, i.e., cancer, diabetes and CVD; Biological age acceleration: difference between the individual’s biological age estimate, *SurvMetaboAge*, and their chronological age.

Two sensitivity analyses for the discriminatory ability of the risk scores are presented in the Supplementary Material Figure 11. Firstly, Cox models were refit for the participants of age 70 and older and it was found that BAA is still informative for the survival, with just a slightly lower C-statistic than in the whole cohort, 0.681 and 0.690 for men and women, respectively. The discriminatory ability of common risk factors and prevalent conditions in this group is lower when compared to the entire validation cohort, demonstrating the importance of the NMR metabolite score. Secondly, models were fit on a validation cohort subset excluding statin medication users, where the results were very similar to the main analysis.

To better understand the added value of NMR metabolite biomarker score, we analysed the reclassification of the model based on the biological age acceleration with respect to the model based on common risk factors and prevalent disease for 5-year mortality risk (see Supplementary Table 5 for the details). Risk category-based net reclassification improvement (NRI) based on four risk categories (*<* 0.5%, 0.5–5%, 5–10%, *>* 10%) was 12.6% (95% CI 8.8%–16.1%), comprising 5.6% (95% CI 3.0%–7.2%) for survivors and 7.0% (95% CI 4.5%–10.1%) for non-survivors. The risk difference based (category-free) NRI was 23.7% (95% CI 13.8%–30.8%), comprising 18.0% (95% CI 12.9%–22.6%) for survivors and 5.7% (95% CI *−*0.1%–10.4%) for non-survivors. Note that the category-free NRI demonstrates that the the risk estimation is particularly improved for participants with lower probabilities of mortality, which is crucial for a validation cohort with a comparatively short follow-up and low number of deaths (2% in contrast to 18% in the development cohort).

To directly compare biological and chronological age as predictors for mortality, we present ROC curves and area under ROC curve (AUC) values for 5-year mortality based on logistic regression model for three same-age groups (<45, 45–64 and *≥* 65 years) in the validation set, see Figure 6. We can see that unlike the chronological age, the biological age is a good predictor of mortality in each of the same chronological age strata. The most striking advantage of the biological age as a mortality predictor can be observed for younger people (<45 years old), where the chronological age is not informative (AUC 55.76%, 95% CI 50.74%–60.78%), while biological age attains a relatively high discriminatory power (AUC 67.30%, 95% CI 61.75%–72.86%).

**Figure 6:**
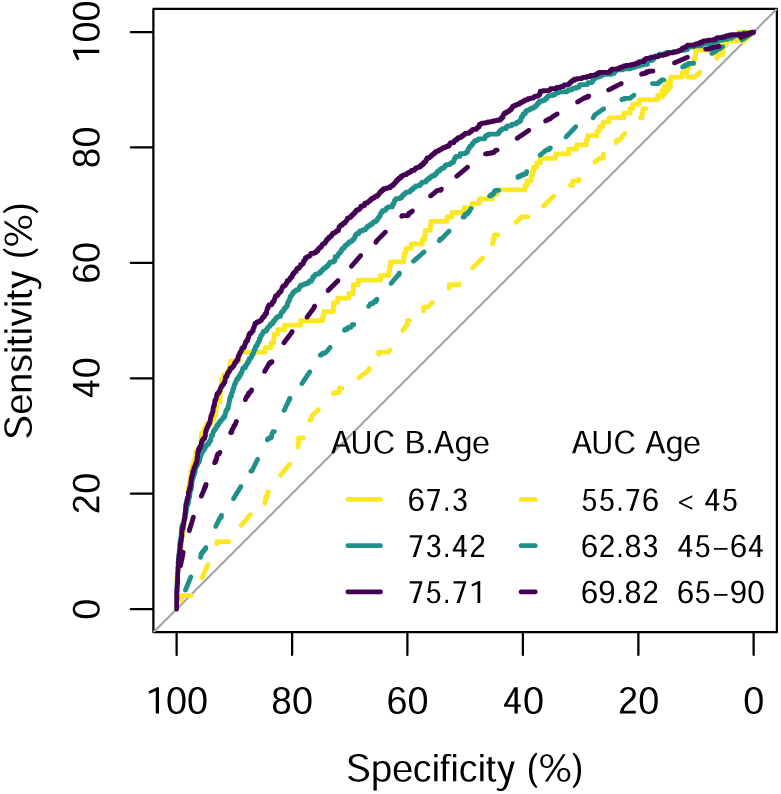
ROC curves and area under ROC curves (AUC) values for 5-year mortality risk. Comparing logistic regression models based on chronological age (Age) and biological age (B.Age), validation cohort stratified by age groups <45, 45–64 and 64–90 years at the joining of the biobank.

Finally, we present the effects (in years) of the common risk factors on the biological age acceleration estimate. Namely, we linearly regress BAA (separately for men and women) on the terms used in the biological age survival model, excluding the metabolite score and including an additional term for statin medication use (Figure 7). First, note that the risk factors explain only about half of the biological age variation as demonstrated by regression *R*^2^. Smoking has the largest effect, being associated with 7.2 (5.6) years increase in biological age for men (women).Diabetes and cancer have the next largest effects. Interestingly, the effects of having the two diseases simultaneously cumulate for women, but not for men, as shown by the values of the interaction term. The effects of cardiovascular conditions (CVD) and BMI>35 are smaller, adding 1-1.5 years to the biological age estimate. The comparatively small CVD effect is probably related to the wide definition that was used for a prevalent cardiovascular condition. In addition, CVD effect is almost cancelled out by the statin usage for women and less so for the men. Finally, a higher education level is associated with a decrease in biological age, which is more important for men.

**Figure 7:**
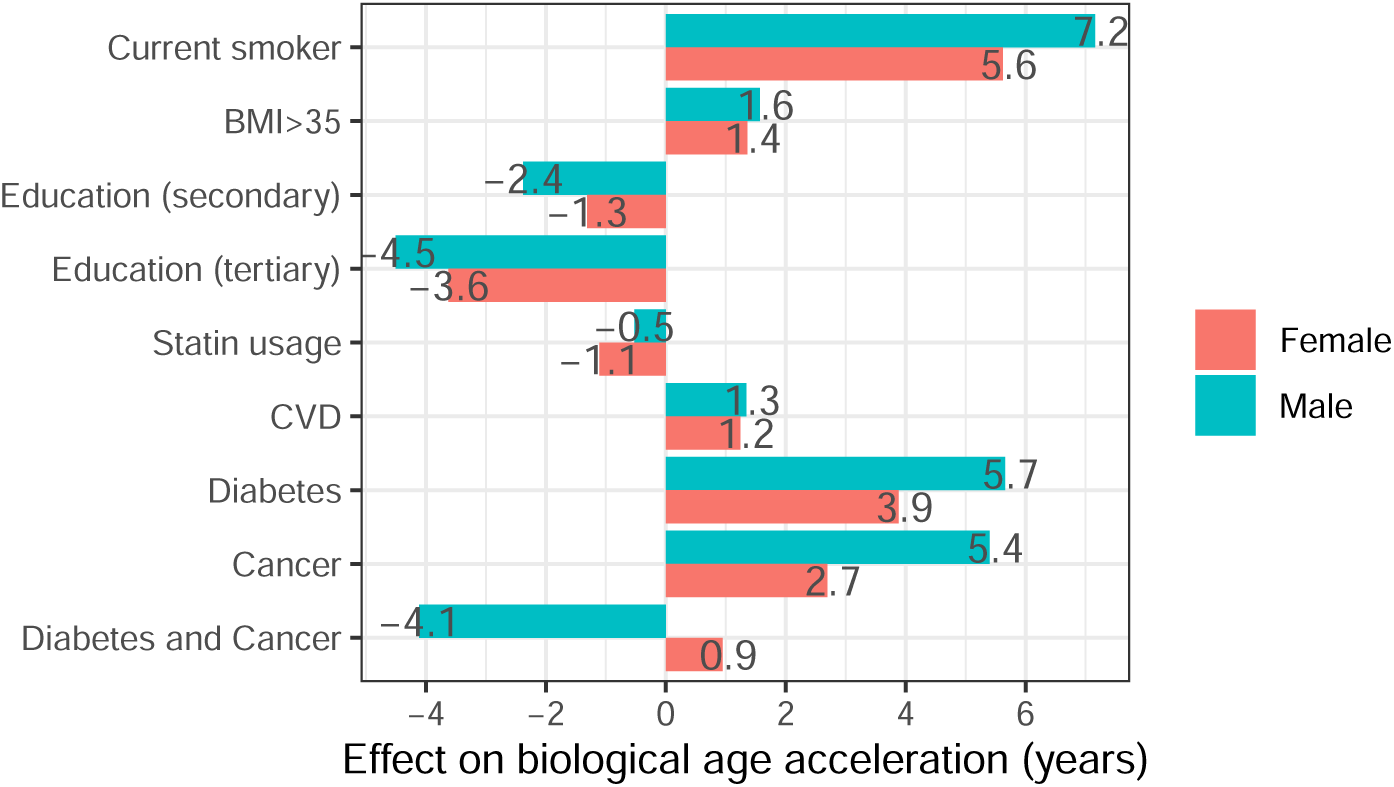
Linear regression model estimates explaining the partial effects of covariates on biological age acceleration in the validation set for men (*n* = 36, 898, *R*^2^ = 49.8%) and women (*n* = 70, 588, *R*^2^ = 45.9%) separately. Effects are estimated with respect to a participant with primary education level, no prevalent disease or statin usage, and having BMI below 35.

## 3 Discussion

We developed a survival-based biological age estimate using a high-throughput NMR metabolomics platform, common risk factors (BMI, education level, smoking) and prevalent disease available for participants of the first cohort (2002–2017, *n* = 31, 358) of Estonian Biobank recruitment. We identified 17 NMR metabolites independently associated with overall mortality and showed that the associations are consistent in the validation set of the second cohort (2018 onwards, *n* = 118, 649) of the Estonian Biobank. The resulting NMR metabolite biomarker score is highly associated with 10-year mortality and stratifies the mortality risk well in the validation cohort.

We defined biological age, *SurvMetaboAge*, as the age where person’s survival probability conditional on his or her covariates is equal to that of the cohorts’ average. The resulting biological age estimate is superior to chronological age for 5-year mortality prediction as demonstrated by higher AUC values in same-chronological-age groups. One year of the biological age acceleration is associated with an important 12% and 17% increase in mortality hazards for men and women, respectively.

Biological age acceleration encompasses all the covariate information in a single and easily interpretable measure while attaining the same discriminatory ability with respect to the survival time. Biological age acceleration estimate also provides an important increase in participant’s mortality risk classification accuracy (23% increase in the category-free NRI) when compared to the model based solely on common risk factors and prevalent disease.

There are several benefits of our model when compared to other biological age models published previously. Firstly, it is trained on mortality (second generation clock) instead of chronological age (first generation clock) as an outcome. The first generation clocks are based on an assumption that the association between biomarkers and chronological age directly informs ageing rates which is not necessarily true and is not testable in a cross-sectional setting [45]. It has been previously noted that the first generation clocks are less predictive for mortality and health-span than the ones directly trained on the survival [33], [43], [24].

Secondly, our biological age estimate is easy to interpret since it is expressed in yearly units instead of hazard ratios or survival probabilities. Thus it can be used not only as a candidate measure of risk stratification in further studies related to health outcomes, but also for health counselling. For example, we found that for women smoking is associated with 5.6 years increase in their biological age, which is a measure easier to communicate than a hazard ratio exp(0.636) = 1.89 following form the survival model.

Thirdly, our biological age estimate is derived directly from a parametric survival model in the discovery cohort without making additional assumptions about the relationship between the hazards and age or the reference population. We shortly comment on the similarities and differences of our method compared to other 2nd generation biological age calculation methods. Our method is the most similar to *PhenoAge*, the first step of the *DNAm PhenoAge* model [27]. Namely, their ‘phenotypic age’ is defined as the age for which the 10-year survival probability of parametric proportional hazards model conditional on selected biomarker values is the same as for a univariate model with chronological age as a single covariate. Our model differs in the sense that instead of time-on-study, we use age as a timescale that is more appropriate for epidemiological studies [46]. *DNAm GrimAge* uses a more complicated method by first associating the biomarkers of interest with DNA methylation rates, and next using the predicted biomarker values (surrogate biomarkers) as covariates in penalized Cox model for overall mortality. Finally, Cox model estimates are linearly transformed to age by forcing the mean and variance of the estimates to be equal to the sample mean and sample variance of the chronological age. In contrast, *Bortz et al.* use a non-parametric Cox regression to associate biomarkers with mortality and assume, following previous work on longevity analysis from the developed world [17], that one unit change in Cox model log hazard ratio results in 10-year deviation in the life expectancy. This method is appealing by its simplicity, however, the question can be posed on its accuracy in a particular population and time cohort. Finally, *AccelerAge* [44] is based on accelerated failure time models and individual’s mean residual lifetime (MRL) conditional on their biomarker profile. As their biological age is a function of MRL and not a function of survival probability, it can be calibrated using the life tables of the reference population. More precisely, the biological age is defined as the age in the reference population where the unconditional MRL equals an individual’s MRL. As noted by the authors, identifying a reference population might not be straightforward, or an appropriate population life table to estimate MRL might not be available, in which case calibration to the discovery sample can be appropriate. To summarise, when choosing the methodology for biological age calculation, one should consider not only the type of outcome for model training, but also the nature of the cohort and reference population of the study.

Finally, our biological age estimate achieves a good performance in the context of omics-based biological age clocks. Currently, epigenetic clocks (DNAm clocks) are considered among the most promising biological age predictors [20]. In comparative studies [13], [33], [48], [24] *DNAm GrimAge* [29] has been shown to have the highest association with overall mortality, with hazard ratios (per SD increase) between 1.76 and 2.10 independent of chronological age. Our biological age acceleration achieves an association of a similar degree, 1.93 (1.94) for females (males) in the validation set. We found that one year of biological age acceleration was associated with HR 1.12 and HR 1.17 increase in mortality hazards for men and women, respectively. This effect size is comparable and even larger to the effects reported for the best epigenetic clocks available, such as *DNAm GrimAge* HR 1.12 (validation meta analysis) [29], *PhenoAge* HR 1.09 [27] and notably larger than for metabolomics-based ageing clocks: *MetaboAge* HR 1.022–1.036 [47], *MileAge* HR 1.04 [35] or *Lau et al.* HR 1.027–1.056 [26] (for different models and validation sets).

Previous research with both targeted NMR-measured [2], [36] and untargeted metabolite platforms [23], [18], [51] has shown sex differences in metabolite profiles. For example, lipid concentrations (particularly triglycerides) in VLDL are higher among adult males while the levels of HDL and LDL cholesterol, apolipoprotein-B, and inflammatory glycoprotein acetyls, are higher among adult females [2]. To account for these differences, we stratified our NMR metabolite biomarker score and biological age model by sex. We observed that the estimate of biological age acceleration had a weaker association with mortality (smaller HR estimate per one year change in biological age acceleration) for women when compared to men despite a larger sample size used to train the models. In addition, the partial effects of common risk factors and prevalent disease on BAA were larger for men, except for simultaneous presence of prevalent diabetes and cancer, and statin usage. Reasons for these differences might be a lower prevalence of baseline risks for the females in EstBB cohort and the inherent variation of female metabolite profiles. Namely, previous studies with repeated measurements have shown that female metabolite profile changes were associated with menstrual cycle phase [8], [30], pregnancy [49] and menopause onset independent of age [50]. Although we excluded pregnant women from the study, we did not control for menstrual cycle stage or menopause status, thus the variation of metabolite profiles might have attenuated the strength of the associations for women’s subgroup. Future research for female participants should address reproductive status variables and include longitudinal measurements to evaluate their association with biological age estimates.

We compared the stepwise-selected metabolic biomarkers included in our biological age estimate with the results of the previous studies and the results were mostly consistent. In our previous study in the Estonian Biobank, alpha-1-acid glycosylation was identified as one of four markers associated with all-cause mortality [9]. It has been suggested, that alpha-1-glycoprotein contributes to GlycA NMR signal [31]. GlycA was among top risk factors for mortality in the current analysis, however, disease-specific analyses showed that it is only significant in cancer and CVD mortality. Albumin and citrate are associated with mortality in our earlier and current study. A larger study where Estonian Biobank was also included among 12 cohorts with over 44,000 samples, identified 14 metabolic markers associated with mortality [7]. Our stepwise-selected metabolites have the same effect direction: histidine, valine and albumin are protective and glucose, lactate and glycoprotein acetyls are associated with increased mortality risk. There were no metabolites with opposite effects between the two studies.

Currently, in the largest dataset (250,341 participants of the UK Biobank) where metabolomic ageing score was developed, 54 metabolites were included in the final model based on sparse LASSO selection [53]. The marker with the largest hazard ratio in that study, glycoprotein acetyls, is also among top markers of increased mortality risk in the current study. In addition, top three protective markers from our study were also protective in *Zhang et al.* [53]: unsaturation, S HDL CE and valine. Additionally, albumin and histidine were protective in both studies and creatinine, acetate, acetone, tyrosine, glucose, citrate and lactate were associated with higher risk in both studies. In addition to NMR-based studies, biological age acceleration has been studied using mass spectrometry-based datasets. For example, in a study including both NMR and mass spectrometry-based datasets in 2239 individuals, pathway enrichment was performed on metabolites associated with BAA, suggesting tryptophan, tyrosine and biopterin pathways relating to an ageing hallmark ‘altered intercellular communication’ are associated with increased biological age acceleration [42]. In our results, tyrosine is associated with increased all-cause mortality, and for non-cancer, non-cardiovascular death cause-specific mortality. The mass spectrometry-based analysis also identified other pathways, highlighting the potential for biological interpretation for the increased coverage of this platform.

We compared our results with the atlas of disease-associated metabolites from the UK Biobank, specifically disease-related mortality [19] in the online database [37]. Metabolic traits in our model that are associated with lower mortality risk, unsaturation, S HDL CE, albumin and histidine, are protective for disease-related mortality for cancer (C), metabolic (E), cardiovascular (I) and pulmonary (J) ICD-10 categories in the UKBB-based atlas of metabolite-disease associations. Valine is protective for dementia and Parkinsons disease-related mortality, but associated with higher risk for mortality of metabolic and hypertension-related diagnoses. From the metabolic traits in our model that are associated with higher mortality risk, omega6/omega3 ratio, glycoprotein acetyls, lactate, citrate, glucose and acetone show consistent results with the UKBB results, while DHA%, XS VLDL FC and acetate are protective for disease-related mortality in the UKBB and tyrosine is protective for some disease-related mortality and associated with higher risk for others. Metabolic traits from our model that showed inconsistent results with UKBB data (protective for some, risk for others), also show similar patterns in our disease-specific death cause analyses. For example, valine which is protective in all-cause mortality model, is protective for some disease-related mortality in the UKBB and risk for others, while our disease-specific analyses also reflect cause-specificity. Tyrosine and glucose, also risk-associated metabolites in our all-cause mortality model, but associated with both risk and protective effects for different disease-related mortality in the UKBB, show differences for cause-specific mortality in our results.

However, there are certain limitation to our study. First of all, out-of-sample prediction should be interpreted with caution. It is well established that biobanks are subject to participation bias, for example, there is evidence that participants of UK Biobank are healthier and live longer than individuals of general UK population [10]. Participation bias can be observed also in Estonian Biobank, for example, females and individuals with higher education are overrepresented [34]. Moreover, differences between the two cohorts can be observed, the newest second cohort being in general more healthy than the first cohort. Nevertheless, the cohort differences can be used for validation, notably, our biological age estimate has shown consistent results in out-of-time validation cohort. Secondly, similarly to most other ageing clocks, our NMR metabolite score and biological age model does not provide causal insights into ageing process [43]. However, it provides testable hypothesis on meaningful biological associations which can be further studied using causal statistics methods, such as Mendelian randomization [53] or genetic colocalization [21]. Finally, our selected biomarkers have demonstrated some disease specificity with respect to the mortality that requires further insight. The current study framework can be extended to develop other disease-specific biological age clocks based on particular outcome of interest, such as ‘heart age’, for example.

In conclusion, we proposed a new method for biological age estimation directly related to individual’s survival. By using a well standardized set of biomarkers based on NMR measured metabolites and common risk factors, we acquired a biological age estimate that is strongly associated with overall mortality. Biological age estimates hold a promise for ageing research as they can be used for risk stratification for targeted interventions. Our biological age estimate also has a natural interpretation as it is expresses in yearly units and thus it is especially suitable for individual health counselling.

## 4 Methods

### 4.1 NMR data handling

We considered the participants for whom 249 circulating plasma NMR metabolite biomarkers were quantified at the baseline using Nightingale Health platform (*n* = 207, 480). NMR metabolites in EstBB have been measured in two rounds, 2012 and 2023. 9,457 participants measured in the earlier round were excluded because Nightingale metabolite quantification protocol has been updated in 2020 and it has been advised against analysing data from different platform versions together [4]. After excluding biomarkers with more than 10% missing values, 238 biomarkers were retained for the analysis including 169 directly measured concentrations and 78 derived ratios. Next, directly measured NMR biomarkers were logarithmically transformed and all biomarkers were scaled to *z* scores. Any *z* scores that exceeded the *±*10 standard deviation level were winsorized. Participants with more than 5 missing biomarker values (*n* = 2, 919) were excluded, and the remaining missing metabolite biomarker values were imputed using the *K*-nearest neighbors method using R Bioconductor package *impute* [12] with sex and age as additional covariates.

### 4.2 NMR biomarker score

We selected 30–90 years old participants, excluding women with possible pregnancies since there is evidence that pregnancy initiates substantial changes in metabolomic profiles [49]. The analytic sample sample size was 150,007 comprising 31,358 and 118,649 participants in discovery (recruitment 2002-2017) and validation (recruitment 2018 onwards) samples, respectively. The mortality data was obtained from the Estonian Death Registry as on April 2024.

We used Cox proportional hazards model stratified by sex with age as a timescale to evaluate the association of biomarkers with 10-year overall mortality. For univariate analysis, significance was explored at *α* = 0.05 level with Bonferroni correction, i. e. 0.05/238. As many of the metabolites were highly correlated, to find a subset of biomarkers independently associated with mortality, we employed Cox proportional hazards model with forward-backward selection with 5% false discovery rate (FDR) corrected entrance criterion [3] given by *p*-value threshold

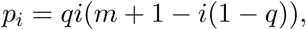

where *q* = 0.05 is the FDR rate, *i* is the model size at the current step, and *m* is the total number of variables considered. The final NMR biomarker score for a particular individual was calculated as their NMR measurements weighted by the respective multivariate Cox model parameter estimates.

### 4.3 Biological age estimation

We define the survival-based biological age as the age where the individual’s current survival probability given his or her covariate profile equals the survival probability of an average individual in the population. By equating the two survival functions, one unconditional and the other conditional on the covariates of interest, we can solve for the survival-based biological age (Figure 8).

**Figure 8:**
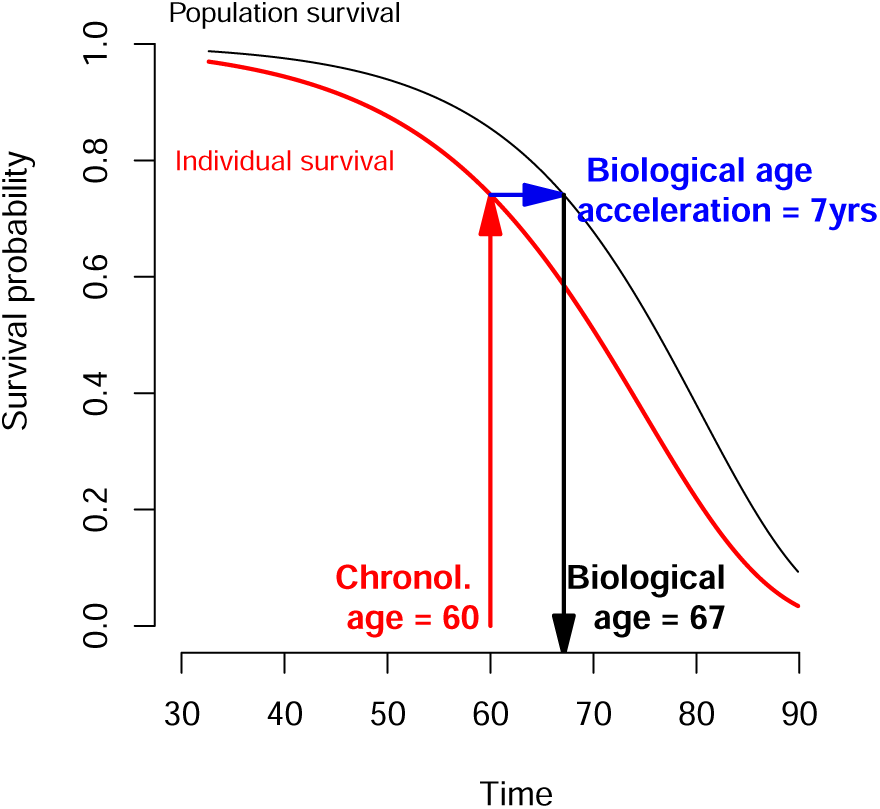
Schematic depiction of survival-based biological age. The black curve represents the population average survival curve estimated without covariates, while the red curve represents individual’s survival curve conditional on his or her covariates. Survival-based biological age is the age where the individual’s survival probability equals the survival probability of an average individual in the population. Biological age acceleration is the difference between the biological and the chronological age. Example for a 60 years old individual with a lower survival probability than average in the population.

We propose to estimate survival probabilities parametrically using Gompertz distribution that is known to approximate the survival of human adults until 90 years of age well [16]. More precisely, Gompertz distribution is given by hazard and survival functions

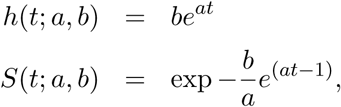

where *a* and *b* are suitable parameters. Let *a_m_* and *b_m_* be the parameters for population average survival *S_p_*(*t*; *a_m_, b_m_*). Let *a_i_*and *b_i_*be the parameters of the *i*-th participant’s survival function *S_i_*(*t*; *a_i_, b_i_*) conditional on his or her *p* biomarker values *x_i_*_1_*, . . ., x_ip_*, where *b_i_* can be expressed as

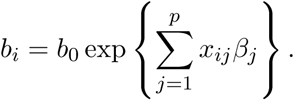

Denote individual’s biological age by *t_B_*, then it can be acquired by solving for *t_B_* in

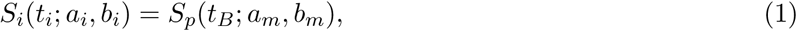

where *t_i_* is the individual’s chronological age. Solving (1) under Gompertz distribution, we have

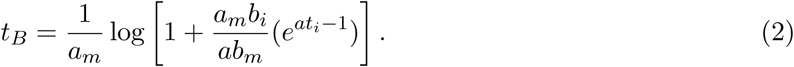

We estimate survival functions and biological age for male and female populations separately, using age as a timescale. We use the standardized NMR metabolite score, common risk factors (BMI, smoking status, education) and prevalent disease status (diabetes, cardiovascular disease and cancer) as covariates in the conditional survival model. We tested for significant interactions of the prevalent disease conditions. We present the correlation with the chronological age using Pearson correlation coefficient. We define the biological age acceleration (BAA) as the difference between individual’s biological and chronological age. We evaluate the ability of BAA to estimate to predict overall mortality in validation cohort by using it as a covariate in Cox proportional hazards model with age as a timescale using Harrell’s C-statistic of concordance as a discrimination criterion. We compare the C-statistics of the BAA model with those of other models containing various alternative sets of covariates. We provide two types of sensitivity analysis: first, we evaluate the models on the validation cohort dataset with participants aged 70 and older, and second, on validation cohort excluding participants using statin medication.

Net reclassification improvement (NRI) for 5-year mortality risk was determined between the reference model based on common risk factors (smoking, BMI, education level and prevalent disease) and the model based on biological age acceleration to examine the added value of NMR metabolite biomarkers to the risk classification. Two versions of NRI were calculated. First, the risk category based NRI was assessed using mortality risk categories 0–0.5%, 0.5–5%, 5–10% and *>* 10%, and second, the risk difference based NRI taking censored participants into account using Kaplan-Meier estimates for the corresponding probabilities [39]. Reclassification was examined separately for survivors and non-survivors, where the net reclassification denotes the sum of the two. R package *nricens* [15] procedures modified for the application of age as a timescale were used for calculations. Finally, we explain the effect of each of the covariates on the biological age acceleration in years by using a linear regression model with biological age acceleration as the dependent variable.

## Ethics and data release

The activities of the EstBB are regulated by the Human Genes Research Act, which was adopted in 2000 specifically for the operations of the EstBB. The data collection and research activities of the Estonian Biobank are performed according to the Estonian Human Genes Research Act (HGRA). Informed consent was obtained from the participants upon joining the biobank, allowing their sample and data to be used for further research. Individual level data analysis in the EstBB was carried out under ethical approval No. 1.1-12/3455 from the Estonian Committee on Bioethics and Human Research (Estonian Ministry of Social Affairs), using data according to release application 6-7/GI/11352 from the Estonian Biobank.

## Data availability

Data used is person-level data of Estonian Biobank participants, which cannot be published by the Estonian law. All data access to the Estonian Biobank’s data must adhere to the informed consent regulations established by the Estonian Committee on Bioethics and Human Research. Estonian Biobank data is of a restricted access. Access is possible on University of Tartu server. Please see https://genomics.ut.ee/en/content/estonian-biobank for the details on the access requests.

## Code availability

Statistical analyses in this study were conducted using R version 4.2.0 (https://www.r-project.org). R code is available from the authors upon request.

## Acknowledgements

The research was conducted using the Estonian Center of Genomics/Roadmap II funded by the Estonian Research Council (project number TT17). K.F. and M.D-V. were supported by the Estonian Research Council grant PRG1197. The work of J.K. was supported by the Estonian Research Council grant PRG1291. This project has received funding from the European Union’s Horizon Europe research and innovation programme under grant agreement No 101060011 “Center for data enriched medicine (TEAMPERMED)”. Views and opinions expressed are however those of the author(s) only and do not necessarily reflect those of the European Union or European Research Executive Agency. Neither the European Union nor the granting authority can be held responsible for them. The metabolomics dataset was produced in collaboration with Nightingale Health. Data analysis was carried out in part in the High-Performance Computing Center of University of Tartu.

## Estonian Biobank research team

Priit Palta, Nele Taba, Erik Abner, Urmo Võsa, Mait Metspalu, Andres Metspalu, Lili Milani, Tõnu Esko

## Supplementary Material A Data preparation

Dataset comprises information from the following sources: Estonian Biobank (EstBB) plasma NMR metabolite sample, EstBB survey data, Estonian Electronic health registry (EHR) data, Estonian Population registry, Estonian Death registry.

### NMR metabolite biomarker dataset

1. EstBB plasma NMR metabolite biomarker sample comprises 215,680 records. The following records were excluded: 2,690 records could not be linked to EHR; 11,969 records created with an older Nightingale platform version in 2012 not advised to be analysed together; 5,709 repeating records on participant level (2nd and any subsequent record for a given participant). 195,319 records remained.
2. From 249 available metabolite biomarkers, 11 biomarkers missing *>* 10% of values were excluded: *XXL VLDL PL pct, XXL VLDL C pct, XXL VLDL CE pct, XXL VLDL FC pct, XXL VLDL TG pct, bOHbutyrate, XL VLDL PL pct, XL VLDL C pct, XL VLDL CE pct, XL VLDL FC pct, XL VLDL TG pct*. 238 biomarkers were remaining for the analysis. Next, the number of missing values on participant level were checked and 245 records with *>* 5 missing biomarker values were excluded. 195,074 records remained.
3. A robust log-scale transform was used for all metabolite biomarkers that were not expressed as percentage ratios (see Supplementary Data 1 for a detailed list).
4. The remaining missing values of metabolites were imputed using K-nearest neighbours method using age and sex as additional covariates.
5. Finally, the outlier impact was limited by winsorizing any metabolite values exceeding *±*10 standard deviations.

### Analytical sample for NMR metabolite mortality score, Table 1

1. 2,730 records with possible pregnancies or 3 months post-partum at the sampling date were excluded. Possible pregnancy was defined as evidence of pregnancy starting (ICD10 codes Z32.1; Z33-Z36) *≤* 365 days before the sample date. 3 months post-partum were identified as pregnancy ending (ICD10 codes O00-O08; O80-O84; Z37) *≤* 90 days before the plasma biomarker sampling date.
2. Participants aged 30 - 90 at the sampling were selected (150,007 records). Records were split in discovery cohort (sampled in 2002-2017, 31,358 records) and validation cohort (sampled 2018 onwards, 118,649 records).
3. Covariates from EstBB survey data were linked (current smoking status and education level). For 37,405 records missing response on education level (99.4% thereof in validation cohort), 36,187 values were imported from the population registry.
4. Covariates from EHR were linked (BMI and prevalent disease status). For BMI, value closest to the sampling date was retrieved and sensibly checked for errors. Prevalent chronic disease status was defined based on the following ICD10 diagnoses codes and time intervals.

a. For diabetes, at least one E10-E14 code before or on the sampling date or at least one O24 code 0 *−* 365 days before the sampling date.
b. Cardiovascular disease: at least one of the following codes: I20-I25, I60-I64, I69, I65-I66, I70, I67.2, I73.9, G45 within 10 years before the sampling date.
c. Cancer: at least one C00-C97 code within 10 years before the sampling date.
5. Covariate of statin medication usage was linked from EHR. ATC codes C10AA01-C10AA08 or C10BX03, first prescribed at least 182 days before the sampling date and last prescribed no more than 365 days before the sampling date constituted definition for stating usage.
6. The outcome for NMR metabolite biomarker score development was defined as 10-year mortality status, i.e., death within 10 years from the sampling, censoring the participants with follow-up below 10 years.
7. Death date and cause was linked form Estonian death registry. Death cause was categorized as Cancer (any C code), Cardiovascular (any I code), Death from external causes (X, Y, V, W) or Other disease (K, J, E, G, N, R, D, F, M, A, B, Q, L, U by the order of the number of outcomes in the training set).

### Analytical sample for biological age modelling

1. As mortality hazards for old ages typically demonstrate deviations from Gompertz law [16], participants older than 90 at the end of the follow-up were excluded from the analysis (1,093).
2. Records with missing covariates of education or BMI were excluded. (1,214 and 10,094 respectively, 10,968 in total).
3. The final analytical sample comprised 137,946 records, 30,460 and 107,486 for discovery and validation cohorts, respectively.
4. For biological age model, we defined the outcome as death within the follow-up. The analysis sample comprised 7,391 death outcomes, 5,129 and 2,262 for discovery and validation sets, respectively.

## Supplementary Material B Comparison of adjusted and unadjusted effects for selected metabolites

We explored more closely the effects of metabolites that showed adverse association with mortality in the univariate (unadjusted) and multivariate (adjusted) Cox models.

### B.1 Metabolites describing fatty acid content

**Supplementary Table 1:** Log-hazard ratio estimates for the three metabolites describing fatty acid content included in the final NMR metabolite score model, *p <* 0.001. Unadjusted: estimates from univariate Cox model; Adj. for one metabolite: estimates from Cox model containing two metabolites; Adj. for all metabolites: estimates from the final NMR metabolite score Cox model containing 17 variables. ns: estimate was not significant, *p >* 0.05. All models are stratified by sex.

| Metabolite | Unadjusted | Adj. for one metabolite: |  |  | Adj. for two | Adj. for all 16 |
| --- | --- | --- | --- | --- | --- | --- |
|  |  | Omega 6:3 | DHA% | Unsaturation | metabolites | metabolites |
| Omega 6:3 | 0.203 |  | 0.208 | 0.136 | 0.336 | 0.226 |
| DHA% | -0.108 | 0.005<sup>ns</sup> |  | 0.175 | 0.477 | 0.262 |
| Unsaturation | -0.201 | -0.159 | -0.353 |  | -0.507 | -0.297 |

We explored DHA% together with the other two metabolites describing fatty acid content (Unsaturation and Omega 6 to 3 ratio). DHA% was a protective factor in the univariate model, but a risk factor in the models adjusted for unsaturation index; unsaturation index and Omega 6 to 3 ratio, and the fully adjusted model, Supplementary Table 1. DHA% effect was not significant when adjusted for Omega 6:3 ratio. Unsaturation and DHA% are highly correlated and exhibit similar size log-hazard ratio estimates with the opposite sign in the adjusted models, however, the correlation pattern is different for the groups of individuals with high and low DHA% values, Supplementary Figure 1. In addition, the correlation between Omega 6 to 3 ratio is dominated by the low DHA% group, the correlation in the high DHA% group being weak. Thus it can be suspected that simultaneous inclusion of the three metabolites in the final model serves a more accurate description of the total fatty acid balance.

### B.2 Metabolites describing lipid balance

We explored M HDL FC pct together with the other two variables describing lipid composition in the NMR score model, XS VLDL FC pct and S HDL CE. M HDL FC pct, the percentage of free cholesterol in medium sized HDL lipids, was a protective factor in the univariate model and when adjusted for XS VLDL FC pct, see Supplemenatry Figure 2. M HDL FC pct is the most strongly correlated with HDL CE (amount of esterified cholesterol in HDL lipids) and M HDL C (amount of total cholesterol in HDL lipids), Supplementary Figure 3. Thus M HDL FC pct might act as a proxy for total HDL cholesterol in our dataset, which is a well known protective biomarker for cardiovascular disease (ref…), hence its protective effect.

**Supplementary Figure 1:**
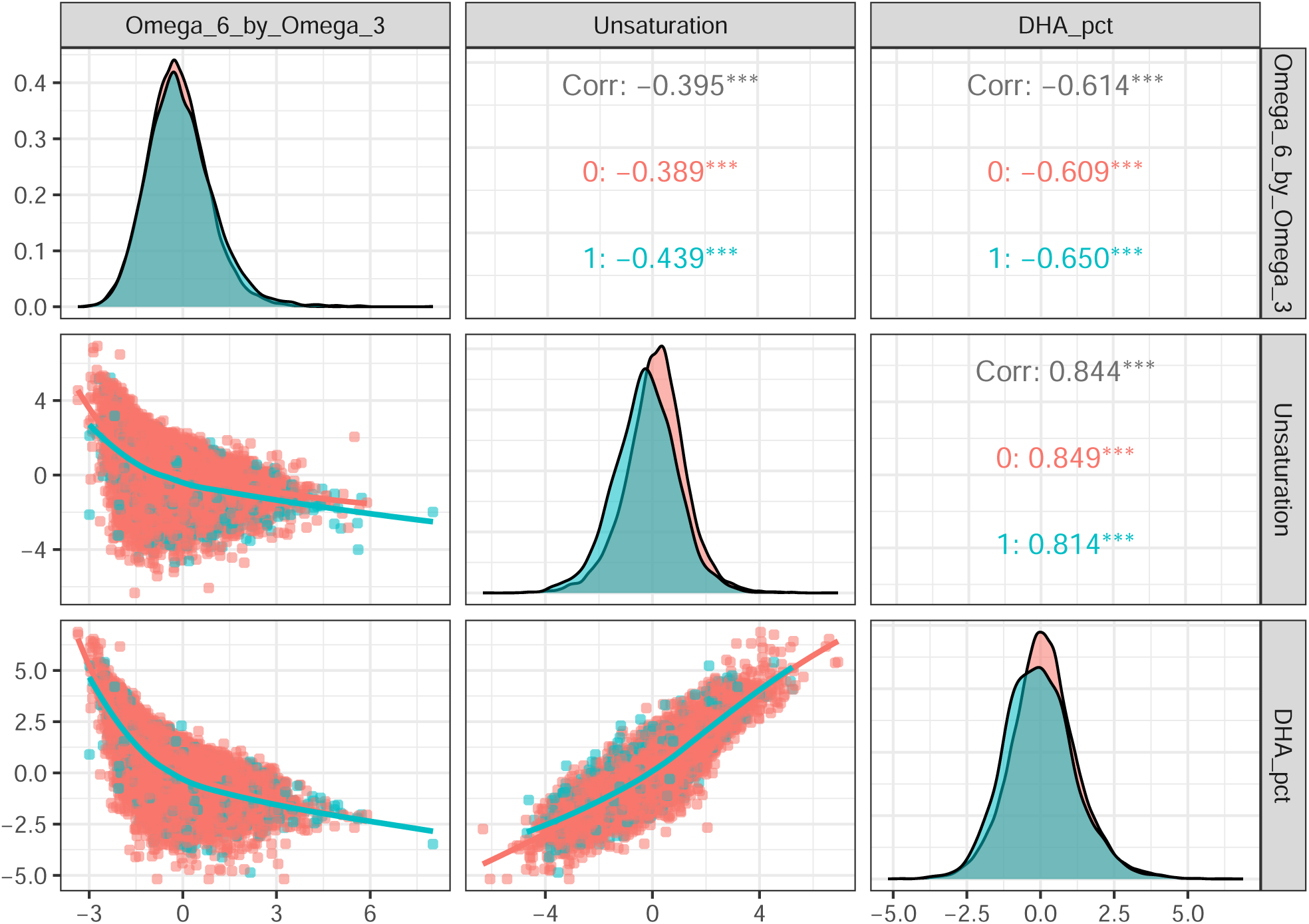
Density plots, Pearson correlation estimates and bivariate correlation plots for metabolites of fatty acid composition included in the final NMR metabolite score model. Gray: ungrouped, red: survived at 10 year follow-up, blue: did not survive at 10 year follow-up.

**Supplementary Figure 2:**
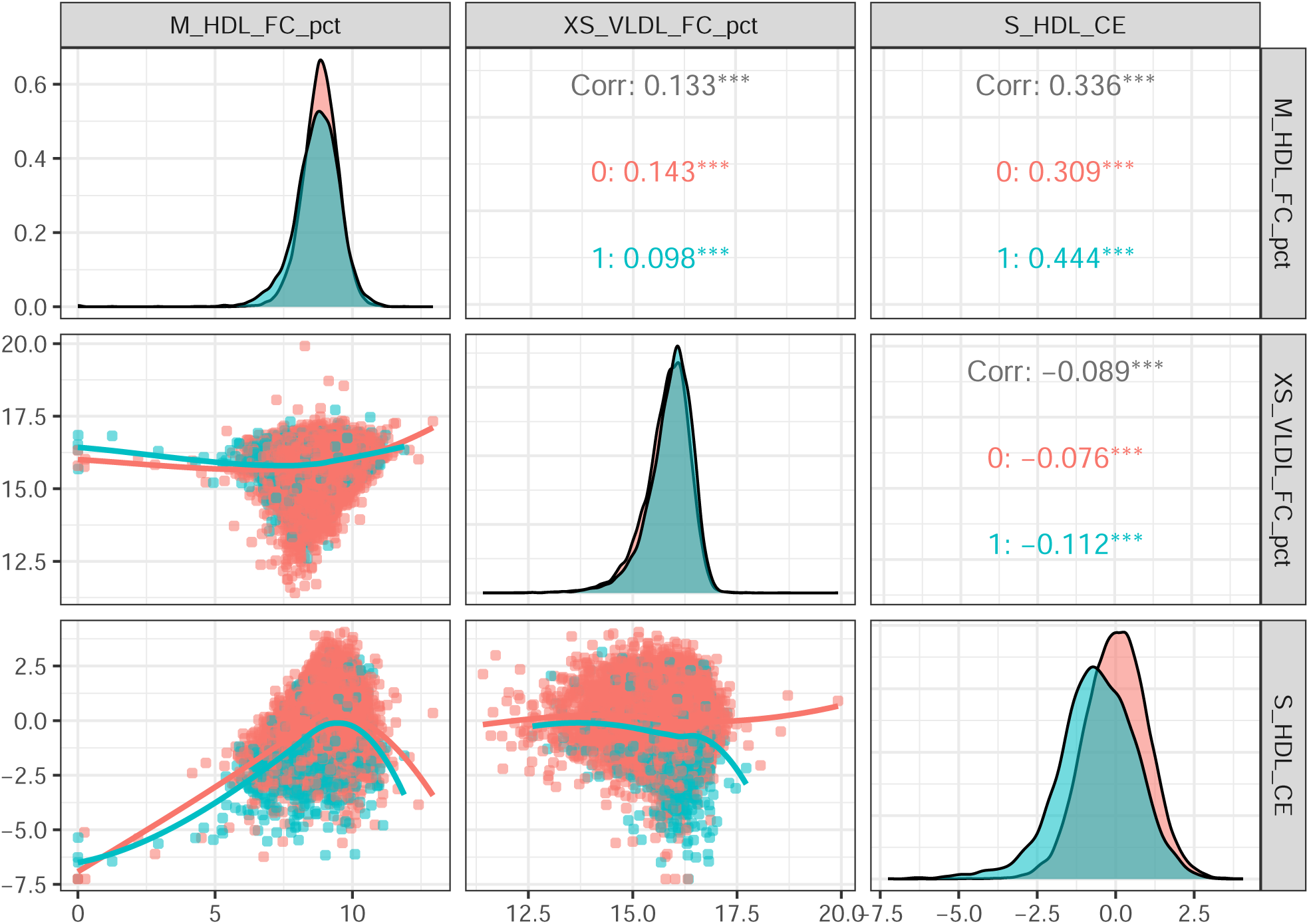
Density plots, Pearson correlation estimates and bivariate correlation plots for metabolites describing lipid composition included in the final NMR metabolite score model. Gray: ungrouped, red: survived at 10 year follow-up, blue: did not survive at 10 year follow-up.

**Supplementary Figure 3:**
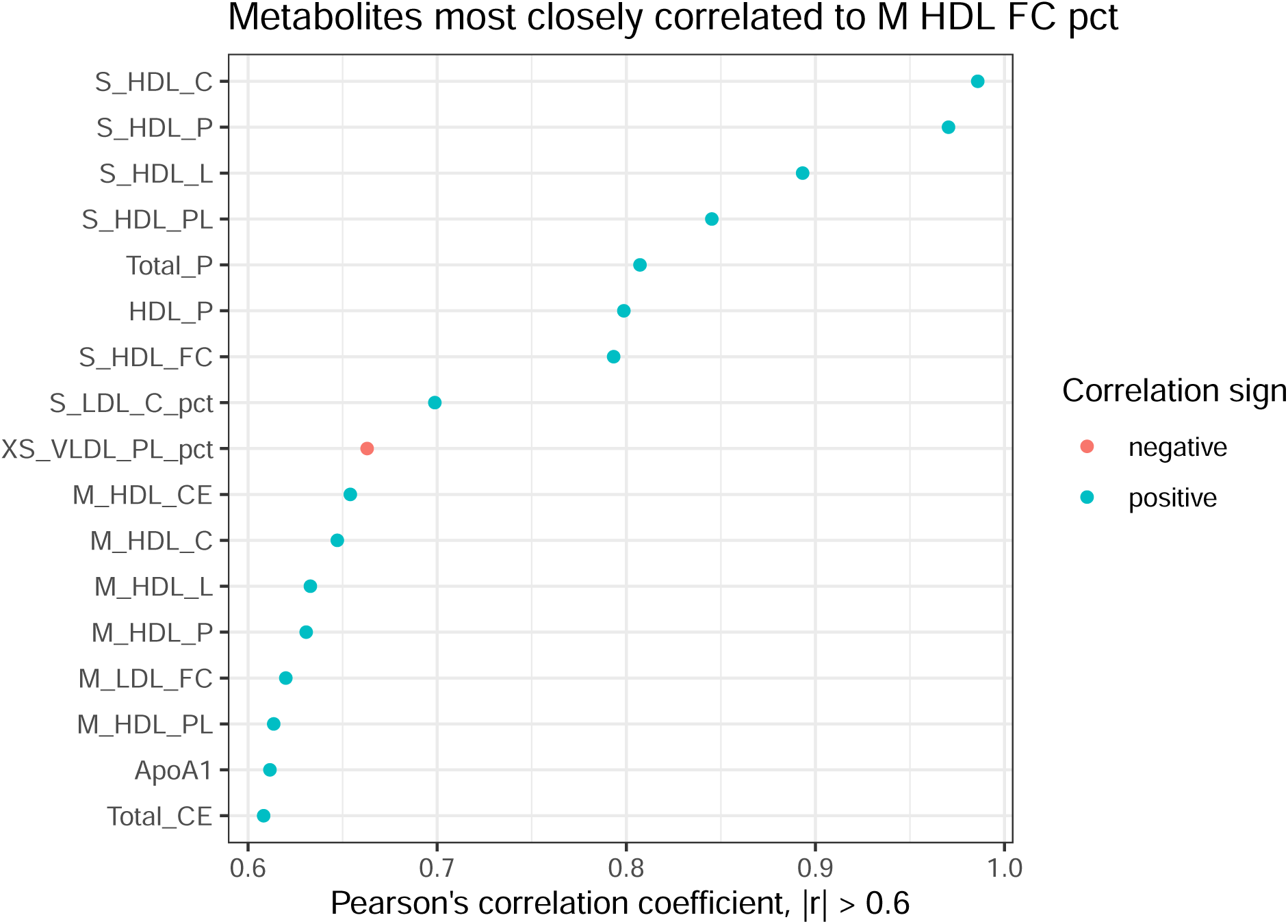
Pearson’s correlation coefficient estimates between M HDL FC pct and other metabolites. Metabolites showing the largest correlations (in absolute values) with M HDL FC pct are displayed.

**Supplementary Table 2:** Log-hazard ratio estimates for the three metabolites describing lipid composition included in the final NMR metabolite score model, *p <* 0.001. Unadjusted: estimates from univariate Cox model; Adj. for one metabolite: estimates from Cox model containing two metabolites; Adj. for all metabolites: estimates from the final NMR metabolite score Cox model containing 17 variables. **: *p <* 0.005. All models are stratified by sex.

| Metabolite | Unadjusted | Adj. for one metabolite: |  |  | Adj. for two metabolites | Adj. for all 16 metabolites |
| --- | --- | --- | --- | --- | --- | --- |
|  |  | S HDL CE | M HDL FC pct | XS VLDL FC pct |  |  |
| S HDL CE | −0.347 |  | −0.369 | −0.347 | −0.366 | −0.167 |
| M HDL FC pct | −0.154 | 0.077** |  | −0.172 | 0.064** | 0.108 |
| XS VLDL FC pct | 0.156 | 0.145 | 0.189 |  | 0.134 | 0.130 |

In contrast, M HDL FC pct was a risk factor in all models adjusting for S HDL CE. Note that S HDL CE is a protective metabolite most strongly associated with mortality (in terms of log-hazard magnitude) among the three metabolites considered. FC (free cholesterol) is unesterified cholesterol which, in excess, can be cytotoxic and contribute to atherosclerosis (ref…). Thus M HDL FC pct in presence of a S HDL CE might act as a risk factor whereas confounding due to lipid structure might occur in the univariate case.

## Supplementary Material C Supplementary Tables and Figures

**Supplementary Figure 4:**
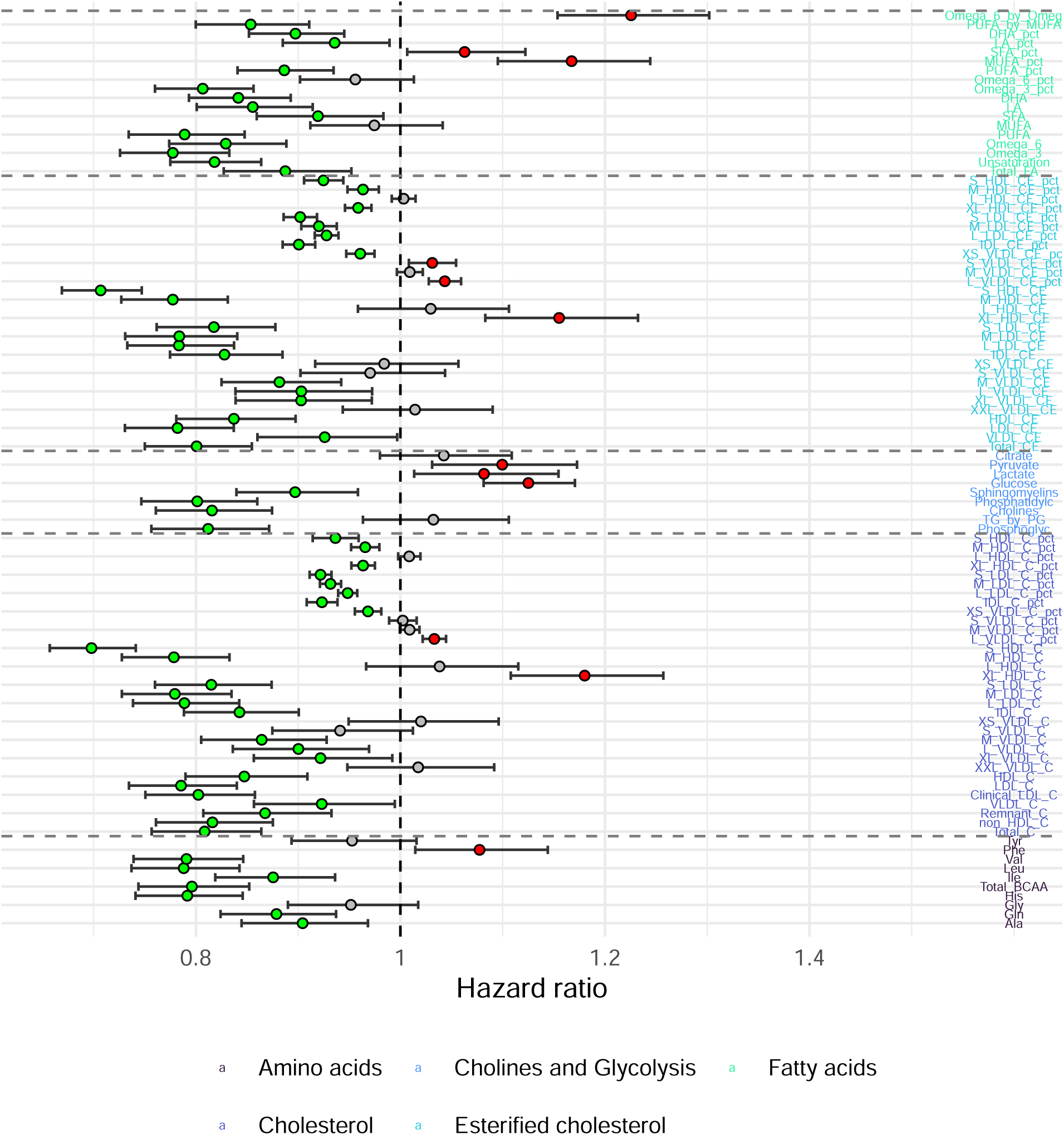
Effects of NMR metabolite biomarkers (part 1) on 10 year mortality, discovery cohort. Univariate Cox model hazard ratios (HR) with the respective Bonferroni adjusted 95% confidence intervals Dashed line represents hazard ratio level equal to 1. Red and green points represent risk and protective effects, respectively, while gray points represent insignificant associations. The colours of metabolite names represent the metabolite classes. FB: fluid balance, GlycA: glycoprotein acetyls, Apo-LP: apolipoproteins. See also Supplementary Data 1 for additional metabolite biomarker statistics (mean, SD, number of missing values).

**Supplementary Figure 5:**
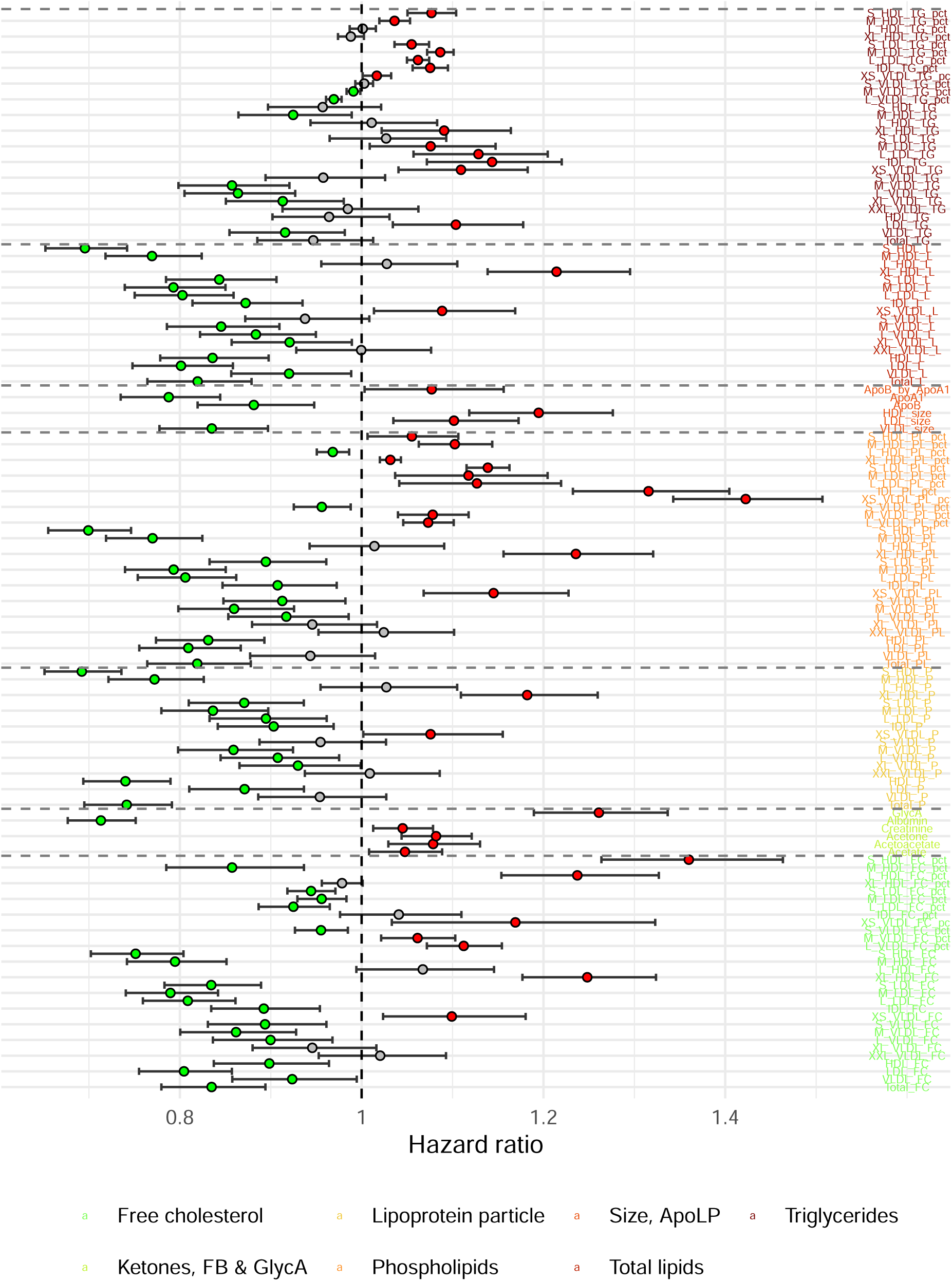
Effects of NMR metabolite biomarkers (part 2) on 10 year mortality, discovery cohort. Univariate Cox model hazard ratios (HR) with the respective Bonferroni adjusted 95% confidence intervals. Dashed line represents hazard ratio level equal to 1. Red and green points represent risk and protective effects, respectively, while gray points represent insignificant associations. The colours of metabolite names represent the metabolite classes. FB: fluid balance, GlycA: glycoprotein acetyls, Apo-LP: apolipoproteins. See also Supplementary Data 1 for additional metabolite biomarker statistics (mean, SD, number of missing values).

**Supplementary Figure 6:**
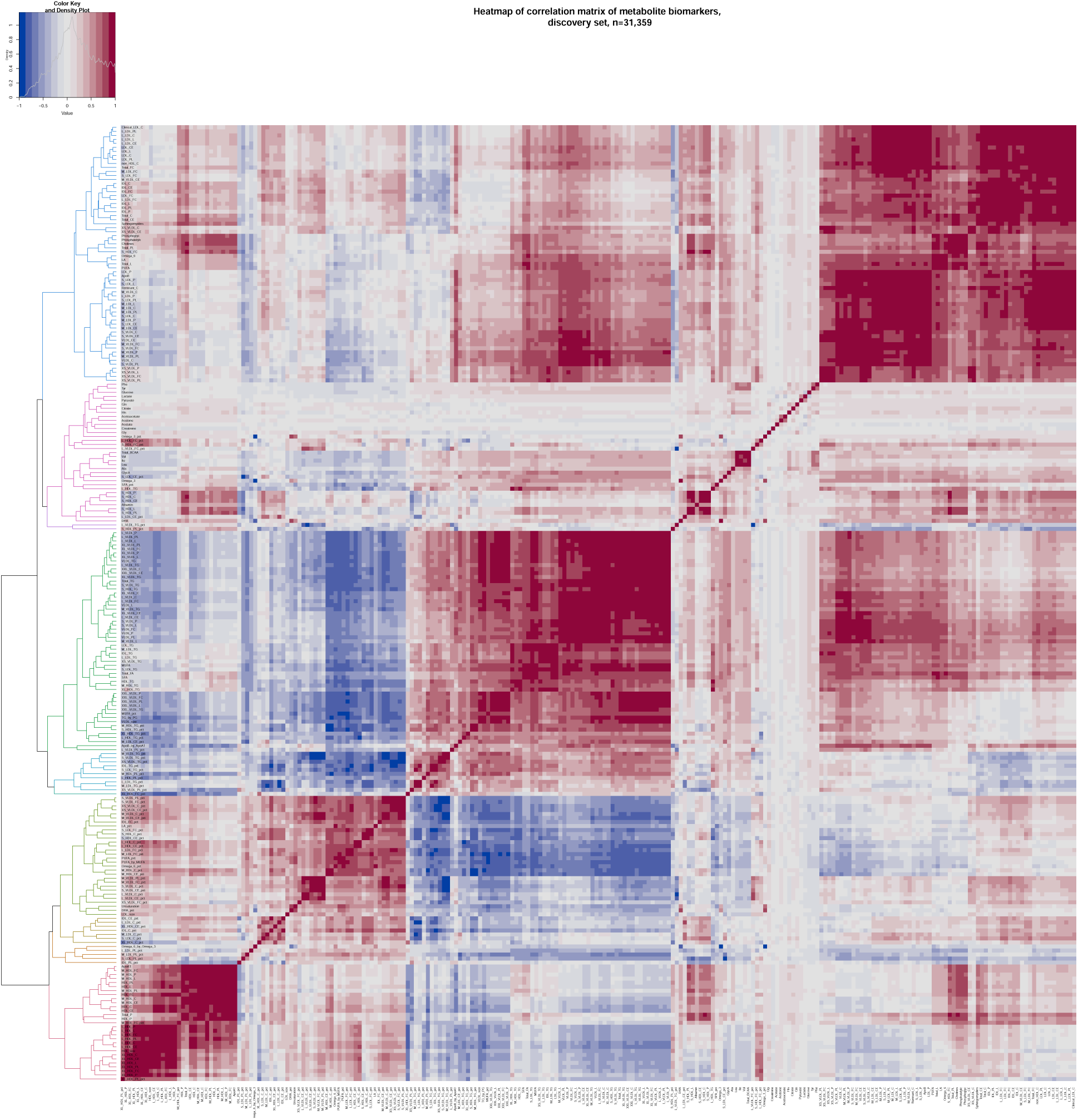
Heatmap and dendrogram representing metabolite biomarker correlations in the discovery set. 10 largest metabolite clusters are coloured in the dendrogram. See file *heatmap_training.pdf* for enhanced readability

**Supplementary Figure 7:**
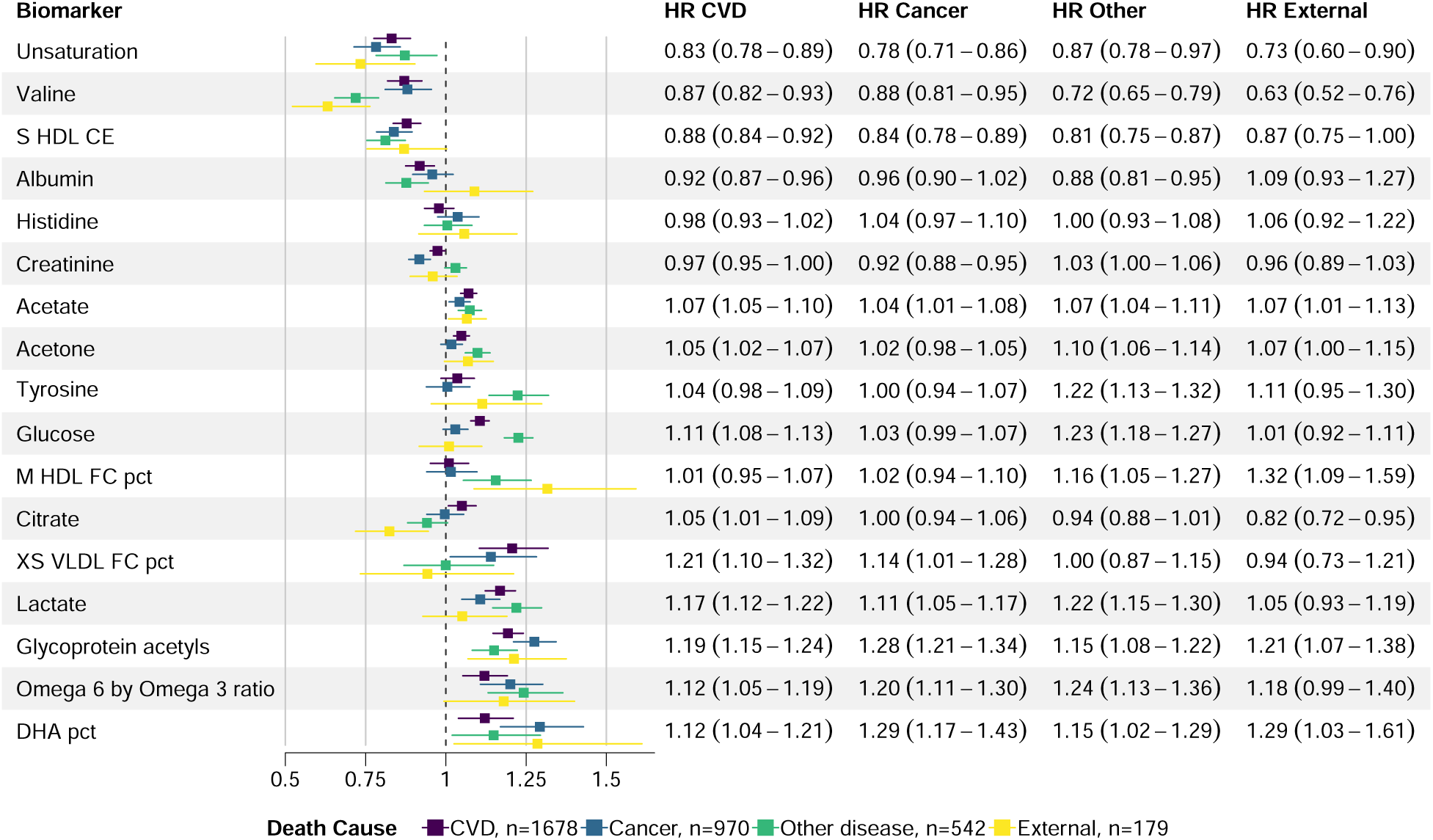
Hazard ratio (HR) estimates (with 95% confidence intervals) per SD change in biomarker value from multivariate Cox models with 17 stepwise-selected biomarkers associated with cause-specific 10-year mortality, discovery cohort. *n* – number of deaths. XS, S, M – lipoprotein size subclass based on the lipoprotein particle diameter; HDL – high density lipoprotein; VLDL – very low density lipoprotein; CE – esterified cholesterol; FC – free cholesterol; pct – percentage measure; CVD – cardiovascular disease.

**Supplementary Figure 8:**
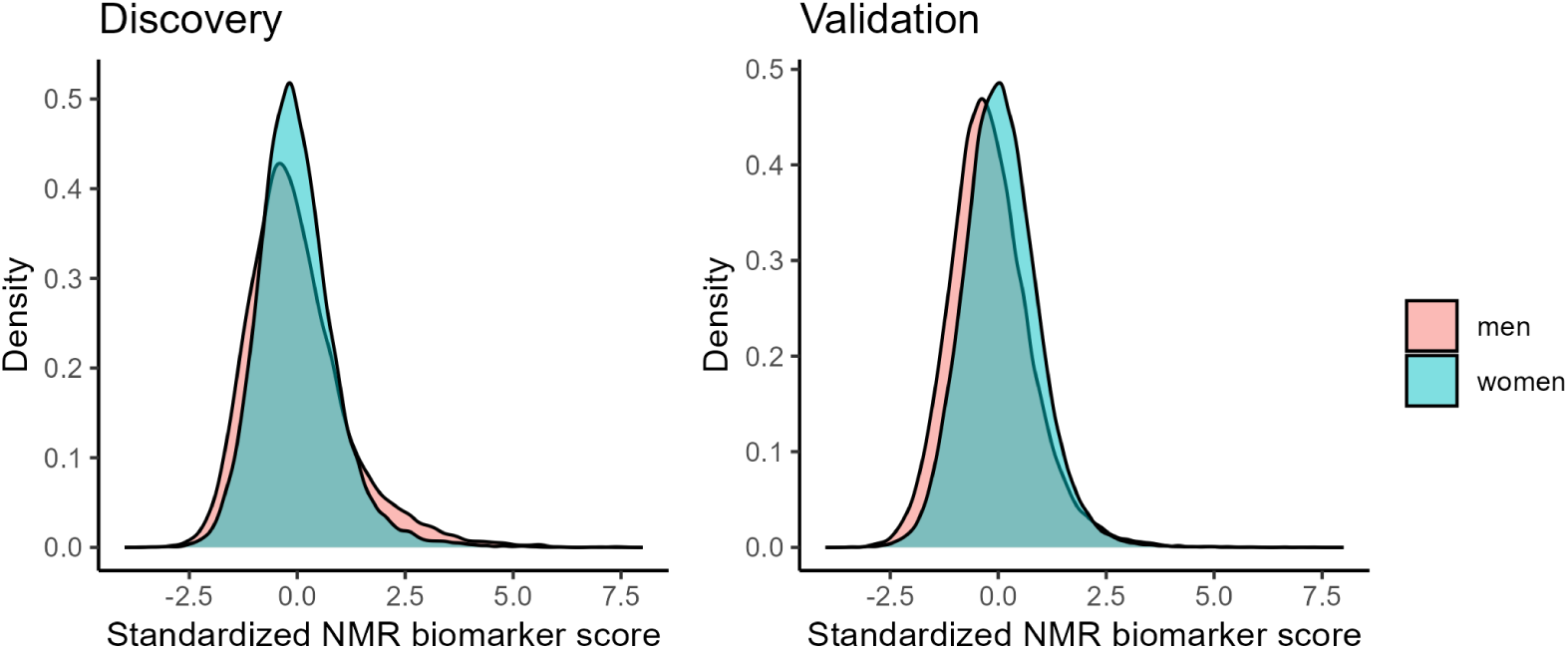
Standardized NMR metabolite biomarker score distribution in discovery and validation sets

**Supplementary Table 3:** Hazard ratios associated with 1 SD increase in NMR metabolite biomarker score related to 10 year mortality in discovery and validation sets, overall and stratified by sex

| Discovery (2002-2017) |  |
| --- | --- |
| Subset | HR (95% CI) |
| All | 1.780 (1.734 – 1.827) |
| Men | 1.921 (1.845 – 2.000) |
| Women | 1.728 (1.667 – 1.791) |
| Validation (2018 onwards) |  |
| Subset | HR (95% CI) |
| All | 1.787 (1.735 – 1.841) |
| Men | 1.819 (1.748 – 1.894) |
| Women | 1.782 (1.701 – 1.867) |

**Supplementary Figure 9:**
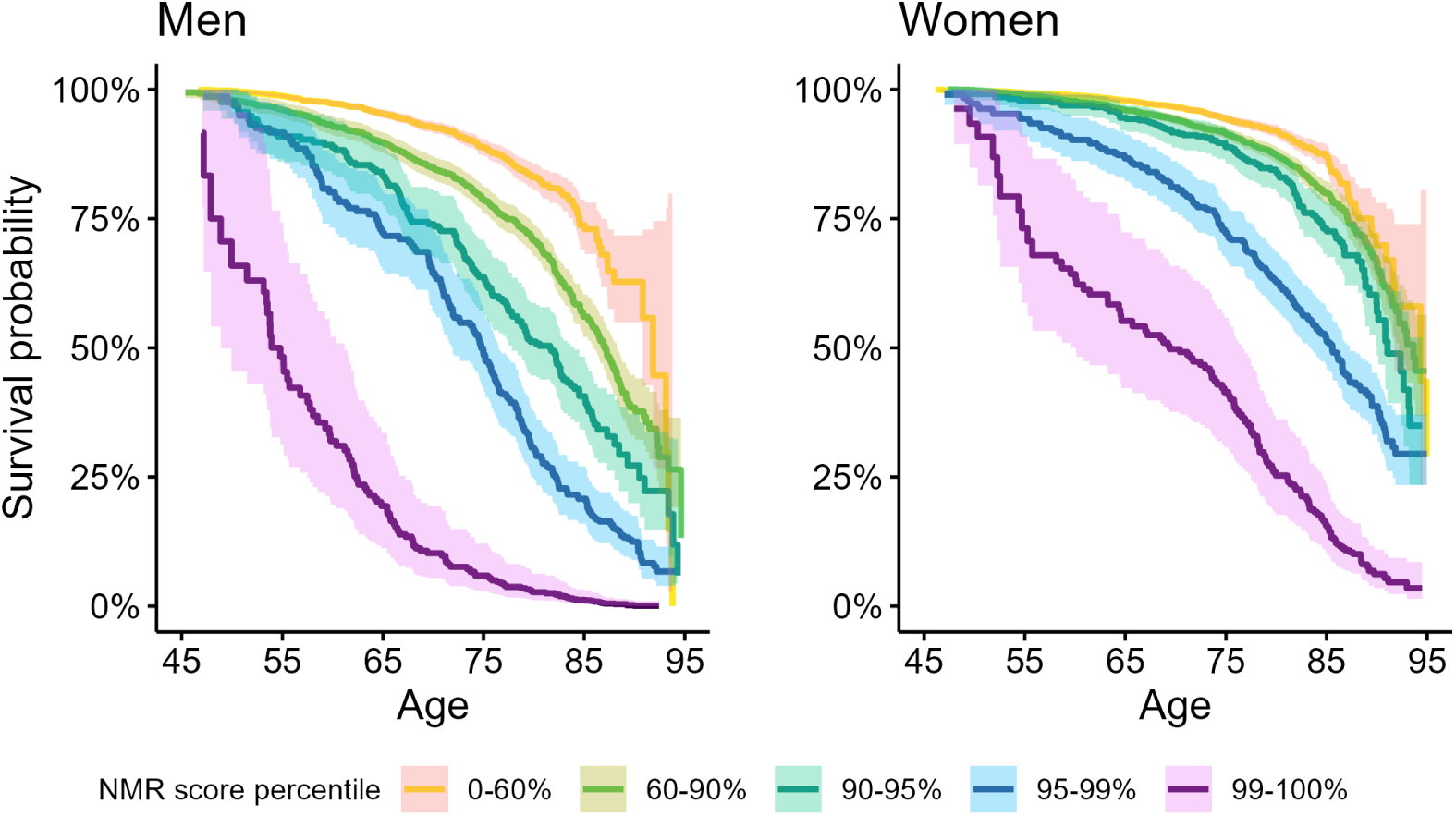
Estimates of overall mortality risk in validation set, stratified by the percentiles of the NMR metabolite biomarker score for participants aged 45 and older. The shaded regions represent 95% point-wise confidence intervals of the survival estimates.

**Supplementary Figure 10:**
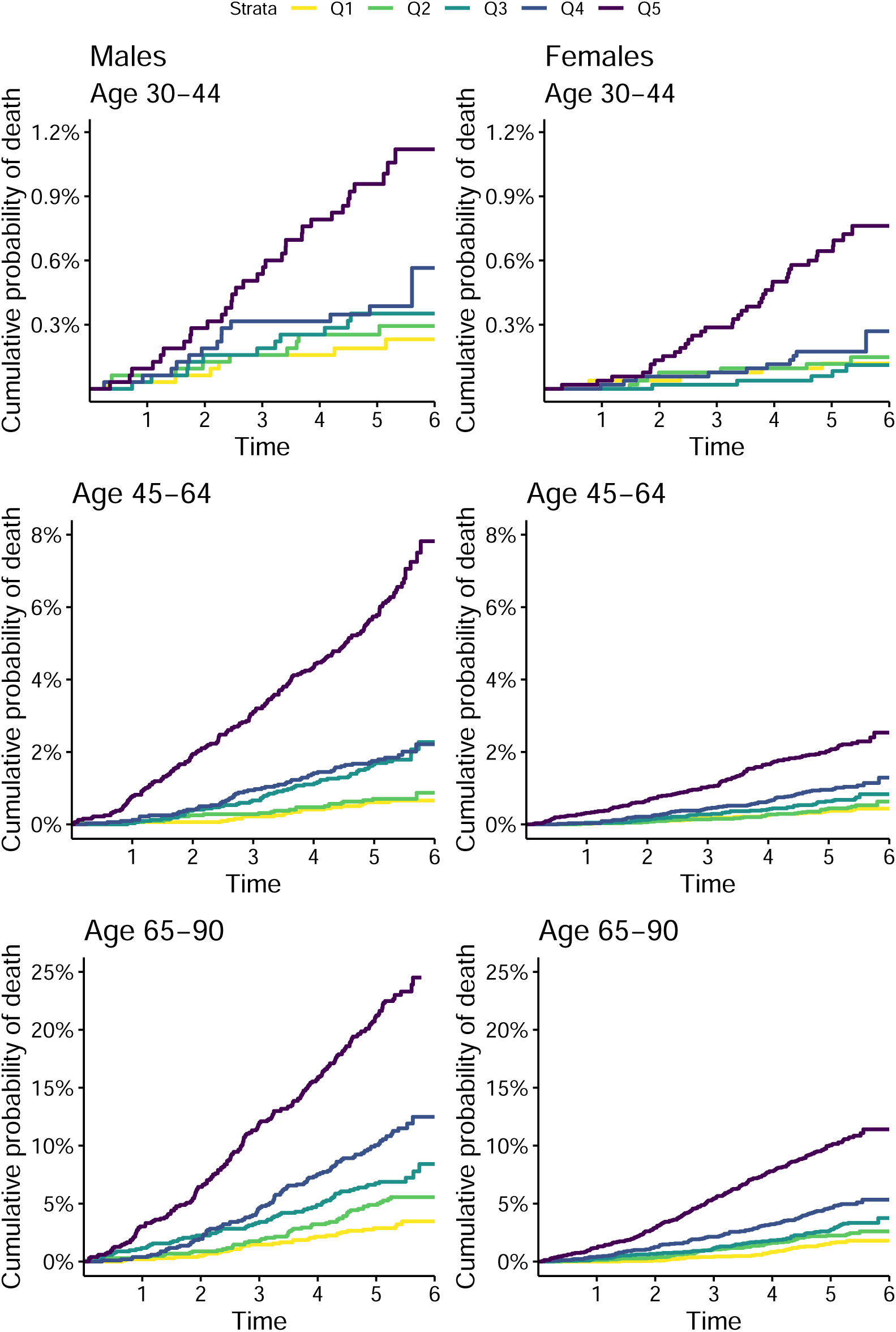
Estimates of overall mortality risk stratified by sex and the quintiles (Q1-Q5) of the NMR metabolite biomarker score compared in the same-age groups 30-44, 45-64 and 65-90. Validation cohort, age timescale.

**Supplementary Table 4:** Parameter estimates for calculating biological age in Equation (2). Estimates are acquired from the Gompertz parametric survival regression models for men and women separately. Standard error estimates are reported in the brackets; for covariates, significance is reported in addition.

|  | Men |  |  | Women |  |  |
| --- | --- | --- | --- | --- | --- | --- |
| $a_m$ | 0.091 | | (0.002) | 0.126 | | (0.002) |
| $b_m$ | -9.700 | | (0.130) | -13.025 | | (0.152) |
| $a$ | 0.081 | | (0.002) | 0.115 | | (0.002) |
| $b_0$ | -9.439 | | (0.213) | -12.765 | | (0.210) |
| NMR metabolite score | 0.434 | *** | (0.016) | 0.475 | *** | (0.018) |
| BMI | 0.003 |  | (0.005) | 0.007 | * | (0.004) |
| Current smoker | 0.543 | *** | (0.046) | 0.636 | *** | (0.058) |
| Education (Secondary) | -0.175 | *** | (0.047) | -0.111 | * | (0.045) |
| Education (Tertiary) | -0.347 | *** | (0.060) | -0.399 | *** | (0.062) |
| Diabetes | 0.302 | *** | (0.062) | 0.325 | *** | (0.055) |
| Cancer | 0.426 | *** | (0.062) | 0.172 | *** | (0.042) |
| Cardiovascular disease | 0.119 | ** | (0.044) | 0.339 | *** | (0.062) |
| Diabetes : Cancer | -0.374 | * | (0.152) | 0.118 |  | (0.130) |
| $N$ | 9, 529 | | | 20, 931 | | |

p < 0.001; \*\* p < 0.01; \* p < 0.05.

**Supplementary Table 5:** Net reclassification improvement (NRI) for the 5-year mortality risk comparing score based on biological age acceleration (BAA) versus the reference score containing common risk factors (smoking, education level, BMI, prevalent chronic disease). (a) Risk category based NRI and the corresponding reclassification tables. NRI is calculated separately for survivors and non-survivors, excluding the censored participants. (b) Risk difference based NRI. Up (down): set of participants who have a higher (lower) risk based on BAA score with respect to the reference score; survivor (nonsurvivor): set of participants who survived (did not survive) during the 5 year follow-up; censored: set of participants, who were followed less than 5 years. The conditional probabilities are estimated with Kaplan-Meier method taking censoring into account. CI: percentile bootstrap estimate of confidence intervals based on 1,000 bootstrap resamples.

|  |  | Risk score based on BAA |  |  |  |
| --- | --- | --- | --- | --- | --- |
|  |  | < 0.5% | 0.5 – 5% | 5 – 10% | 10 – 100% |
| Survivors, $n = 84,352$ | | | | | |
| Reference<br>risk<br>score | < 0.5% | 32,453 | 3,095 | 0 | 0 |
|  | 0.5 – 5% | 7,422 | 35,610 | 1,179 | 127 |
|  | 5 – 10% | 0 | 1,313 | 1,384 | 416 |
|  | 10 – 100% | 0 | 114 | 429 | 810 |
| Reclassification improvement: 0.056, 95% CI 0.030–0.072 |  |  |  |  |  |
| Non-survivors, $n = 1,673$ | | | | | |
| Reference<br>risk<br>score | < 0.5% | 73 | 28 | 0 | 0 |
|  | 0.5 – 5% | 45 | 681 | 101 | 70 |
|  | 5 – 10% | 0 | 73 | 131 | 93 |
|  | 10 – 100% | 0 | 6 | 45 | 327 |
| Reclassification improvement: 0.070, 95% CI 0.045–0.101 |  |  |  |  |  |
| Net reclassification improvement: 0.126, 95% CI 0.088–0.161 |  |  |  |  |  |

(b)
|  | Estimate | 95% CI |
| --- | --- | --- |
| P(down\|survivor) | 0.590 | 0.565–0.613 |
| P(up\|survivor) | 0.410 | 0.387–0.435 |
| P(up\|non-survivor) | 0.529 | 0.495–0.552 |
| P(down\|non-survivor) | 0.472 | 0.448–0.505 |
| NRI survivors | 0.180 | 0.129–0.226 |
| NRI non-survivors | 0.057 | –0.010–0.104 |
| NRI | 0.237 | 0.138–0.308 |
| Survivors | 84,352 |  |
| Non-survivors | 1,673 |  |
| Censored | 21,461 |  |

**Supplementary Figure 11:**
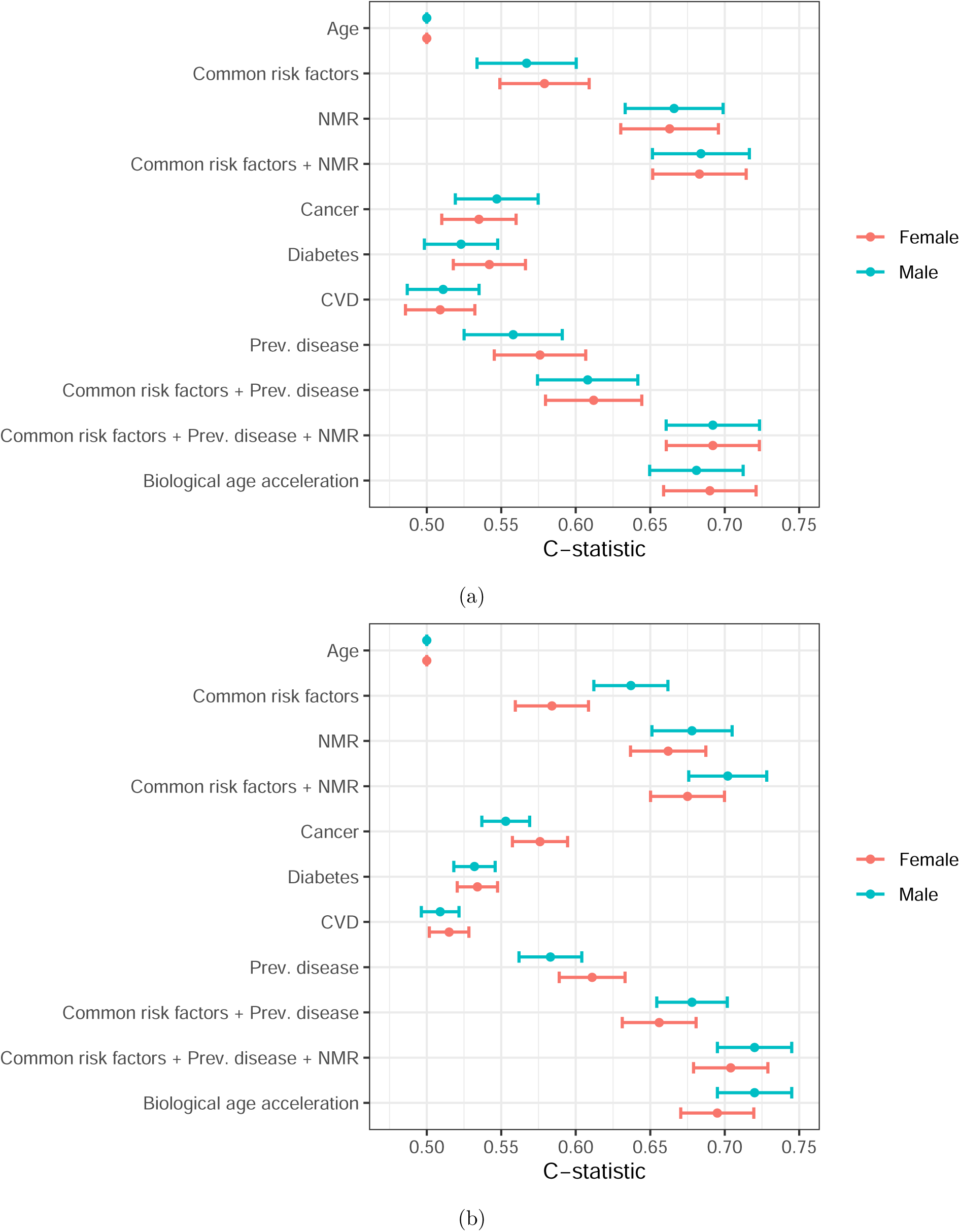
C-statistics of Cox proportional hazards model for survival in the validation cohort. (a): Subset of participants aged *≥* 70, *n* = 5, 997. (b): Subset of the participants excluding statin medication users, *n* = 66, 115.

